# Same household, different choices: variation in health behaviors related to respiratory viruses in Illinois

**DOI:** 10.64898/2026.05.26.26354179

**Authors:** Soren L. Larsen, Junke Yang, Erin M. Haslett, Alyssa Anastasi, Annika Venegas, Lydia Schieleit, Ayesha S. Mahmud, Pamela P. Martinez

## Abstract

While SARS-CoV-2 and influenza continue to place a significant burden on population health, within-household differences in decisions towards vaccination and seeking care across these two pathogens, and across sociodemographic groups, remain largely unexplored. By conducting a household-level survey in Illinois, we found that many individuals made inconsistent decisions about vaccination: among all adults, 29% were vaccinated for only one of COVID-19 or influenza, and among those with children in the home, 39% lived with a child whose influenza or COVID-19 vaccination status differed from their own. A higher proportion of adults were vaccinated against COVID-19 compared to influenza, while the opposite was true for those younger than 18 years old. These differences hold even when accounting for disparities in coverage by age, race/ethnicity, political affiliation, and socioeconomic status. While vaccinated individuals consistently reported wanting to protect themselves or others, those who declined vaccination reported highly heterogeneous reasons ranging from resource constraints to distrust or misconceptions about vaccination. These differences are even more pronounced for COVID-19, with larger partisan gaps and higher refusal driven by safety concerns, lack of trust, or religious reasons than those who decide not to get the influenza vaccine. In contrast to vaccination, the decision to seek medical care when sick showed opposite sociodemographic trends, that are likely attributable to illness severity. Our findings highlight that closing gaps in COVID-19 and influenza vaccination coverage will require an integrative strategy that accounts for diverse motivations, fears, and barriers to access, while addressing social inequalities common to both diseases.

## 2 Introduction

Respiratory pathogens, such as SARS-CoV-2 and influenza, are responsible for a substantial disease burden in the U.S. [1–3], and there are sociodemographic disparities in how this burden is distributed across the population (e.g. [4–6]). Resource access and healthcare utilization were highly heterogeneous during the COVID-19 pandemic, most notably for testing [7], vaccination [8, 9], and treatment [10]. Similar patterns have been reported for influenza, for example with respect to vaccination disparities [11, 12] or access to healthcare during the H1N1 pandemic [13].

Previous studies have also examined vaccination decisions or other healthcare-seeking behaviors for respiratory illnesses, including the role of age, income, race/ethnicity, political affiliation, and other factors ([14– 19]). Some have compared COVID-19 versus influenza vaccination hesitancy or intentions (e.g. [20–22]), while others have examined parents’ behavior in relation to their children, such as parents’ hesitancy around childhood influenza vaccination [23] or COVID-19 vaccination [24–26]. However, little work has been done to comprehensively characterize health-seeking behavior for respiratory diseases - across pathogens, households, and proactive (vaccination) versus reactive (seeking care when sick) behavior - in the same study population. For example, we have limited understanding of whether parents are making the same health-seeking decisions for themselves as for their children across sociodemographic groups, or whether these patterns are the same for COVID-19 versus influenza. Understanding these intersections is key to ensuring equitable access to healthcare and the maximization of public health resources.

To address this gap in knowledge, we fielded a survey in Illinois during the 2024-2025 respiratory disease season, gathering information on disease-related behaviors for COVID-19 and influenza across age, households, and key sociodemographic variables. We found ongoing disparities in the overall reported vaccination coverage across demographic groups, including by age, race/ethnicity, and measures of socioeconomic status. Adults were consistently more likely to be vaccinated for COVID-19 than influenza regardless of demographic group, while the reverse was true for children - resulting in households with mismatched vaccination statuses between adults and children for both pathogens. While we observed similar motivations among those who were vaccinated, reasons for non-vaccination were highly heterogeneous across diseases and age groups. Finally, we found that decisions related to health seeking behavior had opposite trends across groups and appeared to be driven more by illness severity rather than access or willingness to seek care. Overall, differences in proactive and reactive health-seeking behavior are not explained by eligibility, access, or mistrust alone, highlighting the need for more targeted interventions rather than one-size-fits-all strategies.

## 3 Results

Of the unweighted sample of 8, 741 survey respondents aged 19 years or older, 42% [95% CI: 42%, 43%] reported receiving both an influenza vaccine within the 12 months preceding the survey, and a COVID vaccine with at least one booster. Among adults in the survey, 9% [8%, 9%] reported vaccination for influenza only, and 20% [20%, 21%] reported vaccination for COVID-19 only (Figure 1A). Adults with a child in the home were less frequently vaccinated for both diseases than those without children, due to their lower rates of COVID-19 vaccination. Similarly, children, whose vaccination behavior was reported by adult respondents in the home (2, 438), were less frequently vaccinated for both pathogens than adults (30%, [29%, 31%]). Among survey respondents who reported both their own and a child’s vaccination status (n = 2, 380, Table S3), 977 respondents (41%) reported some degree of mismatch between their vaccination status for influenza or COVID-19 and the child’s status. In particular, 20% [19%, 21%] reported being vaccinated for COVID-19 while the child was unvaccinated, and 8% [8%, 9%] reported the same for influenza. In contrast, 16% [15%, 17%] were not vaccinated for influenza but reported a vaccinated child in the home, and 5% [4%, 5%] reported the same for COVID-19. After applying population weights, these coverage estimates remained similar.

**Figure 1:**
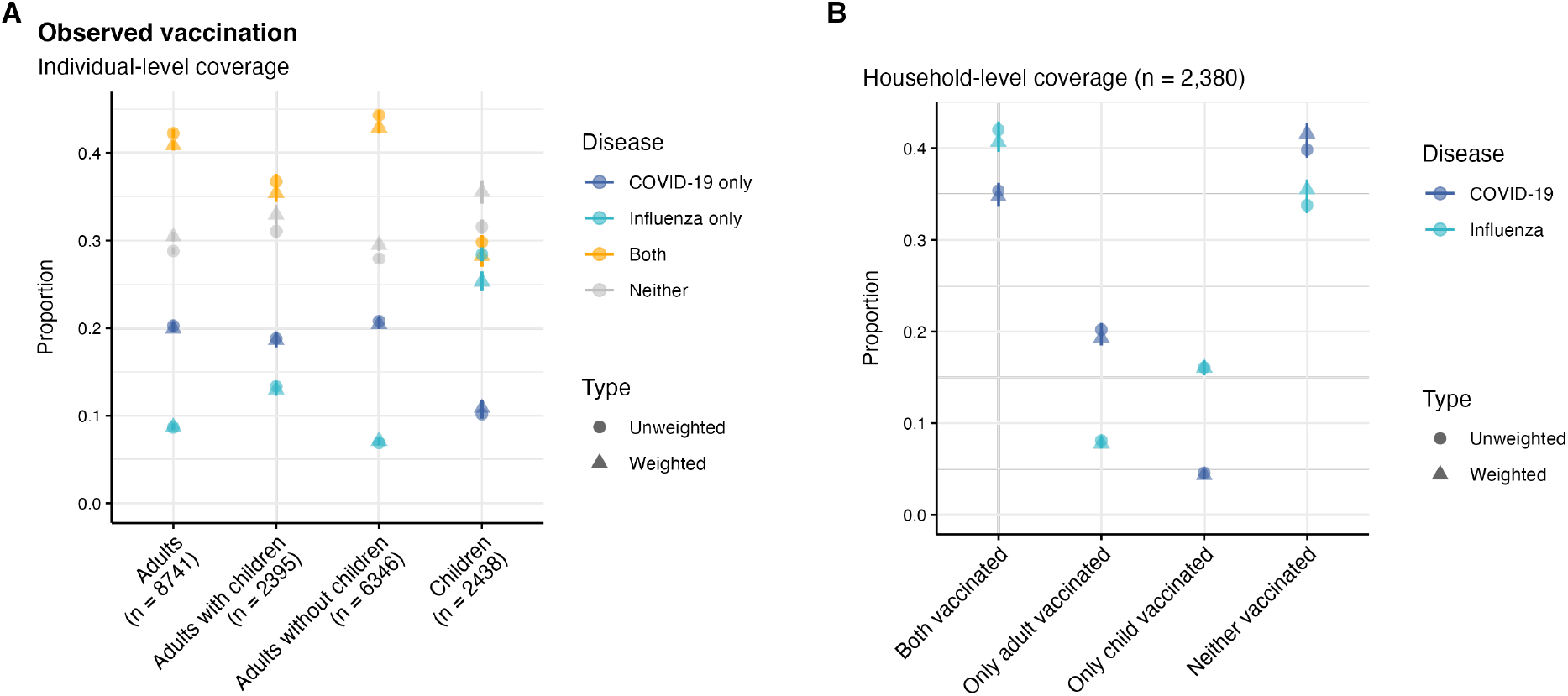
Observed influenza and COVID-19 vaccination coverage in Illinois. Unweighted and weighted vaccination coverage is shown in panel (A) for all adults, adults with children, adults without children, and children of adult respondents. For each group, the total number of respondents is provided in parentheses. In panel (B), the unweighted and weighted vaccination coverage is shown for adults reporting both their own vaccination status and that of a child in the home. For all reported values, 95% confidence intervals are shown. Sample characteristics are provided in Tables S1-S2.

Seeking to understand the factors contributing to these observed coverage patterns, we next disaggregated adult and child vaccination coverage by age, income, education, race/ethnicity, and political affiliation (Figure 2). We find sharply increasing adult vaccination coverage with age. For example, 0.43 ([0.40, 0.47]) of respondents aged 19-29 are vaccinated for COVID-19, rising to 0.89 [0.82, 0.93] among age 80+. We also find that higher income and educational attainment were associated with substantial increases in the proportion vaccinated for both diseases (Figure 2A). Asian participants had the highest vaccination proportions of any racial/ethnic group for both diseases (0.80 [0.76, 0.84] for COVID-19, and 0.60 [0.55, 0.65] for influenza), and Democrats had the highest proportions vaccinated of any political affiliation (0.75 [0.73, 0.77] for COVID-19; 0.62 [0.60, 0.64] for influenza). Despite clear disparities in vaccination coverage between sociodemographic groups, which were qualitatively similar by pathogen, adults consistently reported higher vaccination rates for COVID-19 than influenza, regardless of age, income, education, race/ethnicity, or political affiliation. In contrast, children were consistently reported to have higher influenza coverage than COVID-19, but there were qualitatively similar demographic disparities as those seen for adults (Figure 2B). COVID-19 coverage increased substantially from age 0-5 to age 6-18, with slightly higher coverage among age 6-18 (0.45, [0.42, 0.49]) than those 19-29.

**Figure 2:**
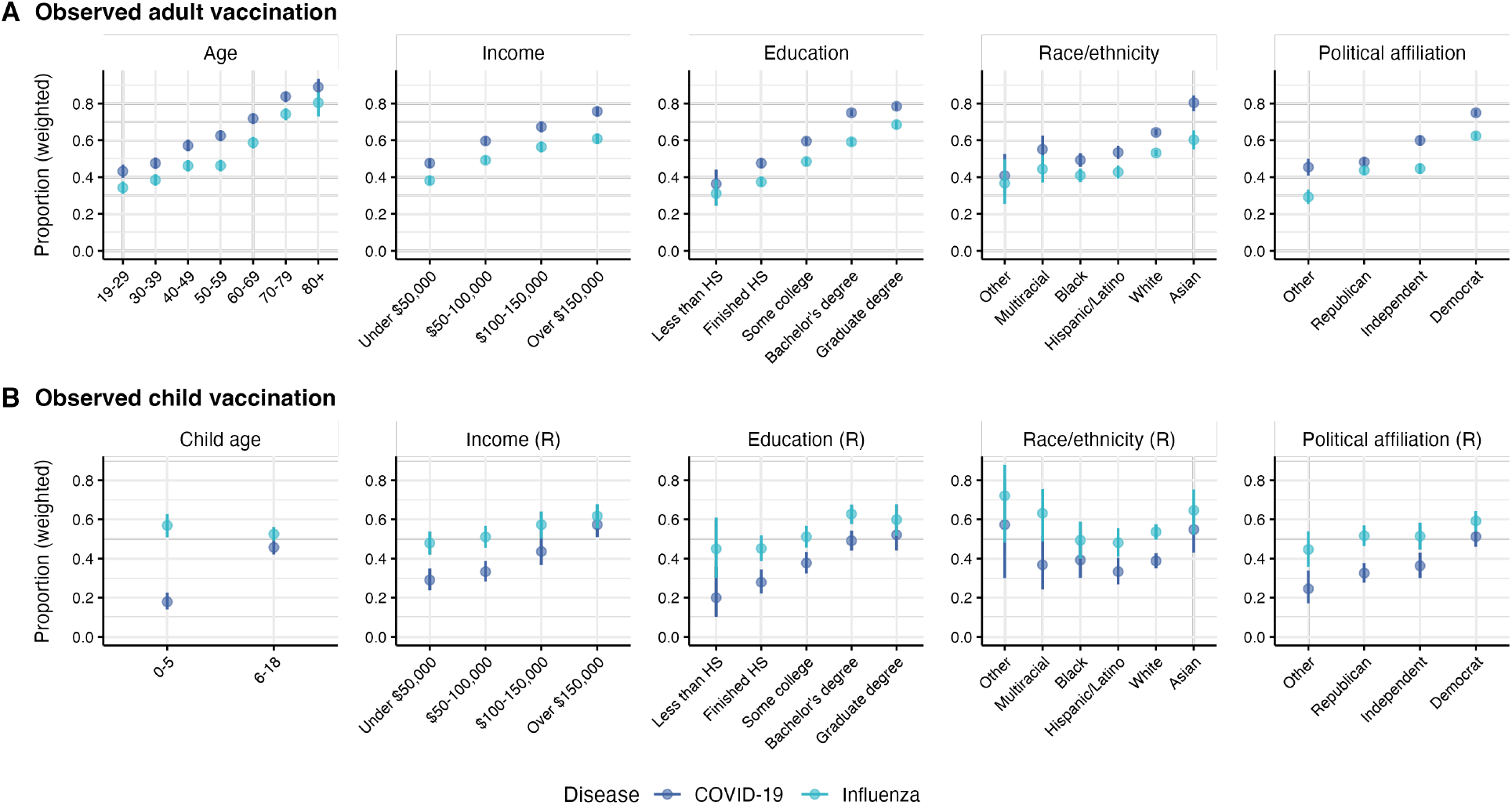
Observed influenza and COVID-19 vaccination coverage in Illinois by sociodemographic group. For adults (*n* = 8, 741 respondents), weighted vaccination coverage is shown in panel (A) by age bands, income, education level, race/ethnicity, and political affiliation. For children of adult respondents (*n* = 2, 438), weighted vaccination coverage is shown in panel (B) by age band of the child, respondent income, respondent education, respondent race/ethnicity, and respondent political affiliation. Weighted 95% confidence intervals are shown for all reported values. Sample characteristics are provided in Tables S1-S2.

Next, we fit weighted logistic regression models to isolate the role of these demographic features (Tables S4, S5, S6, S7), controlling for employment, gender, and survey wave. Predicted vaccination probabilities were computed from these fitted models for each set of sociodemographic characteristics, with all other covariates held at reference categories (Democrat; Unemployed; White; Male; No degree/diploma; income under $50,000 annually; Wave 1; age 19-29 years). Among adults (Figure 3A), age, income, education, race/ethnicity, and political affiliation were all significant predictors of both COVID-19 and influenza vaccination status (Tables S4-S5). The adjusted probabilities of vaccination across age were consistent with the patterns observed in Figure 2, although adjusted gradients across income were notably flatter than in the observed data, with only a 10 percentage-point increase in probability between the lowest and highest income levels for COVID-19, and a 6 percentage-point increase in probability for influenza. Asian respondents remained the most likely to be vaccinated for both pathogens after controlling for other characteristics (*p<* 0.001); similarly, Republicans remained less likely than Democrats to be vaccinated for both pathogens (*p<* 0.001).

**Figure 3:**
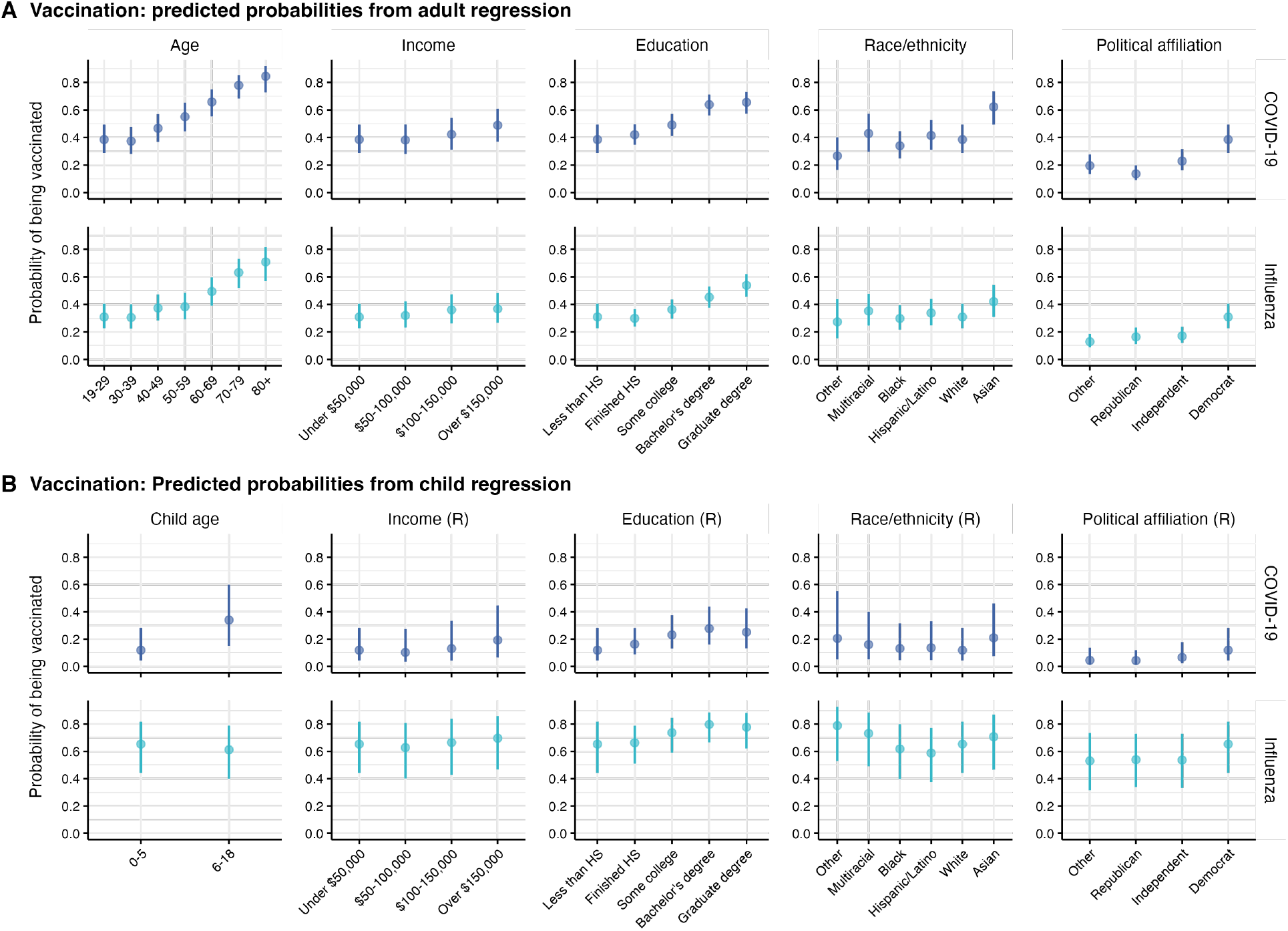
Predicted probability of being vaccinated for influenza or COVID-19 as an adult or child, from four generalized linear models. For adults, the predicted probability is shown by age bands, income, education level, race/ethnicity, and political affiliation. For children, the predicted probability is shown by age band of the child, and respondent income, respondent education, respondent race/ethnicity, and respondent political affiliation. 95% confidence intervals from regression analysis are shown for all reported values. Sample characteristics are provided in Tables S1-S2 and regression parameters/significance levels in Tables S4, S5, S6, S7.

Among children (Figure 3B), the age of the child as well as the adult respondent’s age, income, education, race/ethnicity, and political affiliation were all significantly predictive of COVID-19 vaccination status (Table S6). In contrast, only the adult respondent’s political affiliation and education were significant predictors for influenza, whereas race/ethnicity, income, or respondent/child age were not (Table S7). This suggests weaker disparities in influenza vaccination among children compared to adults. Among adults living with children (*n* = 2, 395; Table S8), who tended to be younger than those living without children (Figure S1), we found that age, income, education, race/ethnicity, and political affiliation remained significant predictors of COVID-19 vaccination status (Figure S2, Table S9). Education, race/ethnicity, and political affiliation were all significant predictors for influenza vaccination, but income was not. Respondent age was also significant for the 80+ group, but there was only one respondent in this category, and therefore should be interpreted with caution (Table S10).

Among adults reporting both their own and a child’s vaccination status (*n* = 2, 380; Table S3), mismatched adult/child vaccination status seen in Figure 1 were observed across sociodemographic groups (Figure S3), and we repeated independent statistical analyses to identify predictors of this behavior (Figure S4, Tables S11-S14). The likelihood of being vaccinated for influenza with an unvaccinated child was predicted by race/ethnicity and respondent age, with Asian respondents (*p* = 0.030) and older adults (*p* = 0.016 for those aged 70-79) being more likely to live in these households. For COVID-19, those with children age 6 − 18 were less likely to be vaccinated with an unvaccinated child (*p <* 0.001); this is consistent with our finding that COVID-19 vaccination coverage was higher for older children. Respondent age was also significant for age 80+, but this reflects a single respondent and should be interpreted with caution. The likelihood of being unvaccinated with a vaccinated child was predicted by respondent age for both influenza and COVID-19, with older individuals less likely to exhibit this particular status. For influenza, political affiliation, race/ethnicity, and child age were also predictive. In particular, Independents (*p* = 0.013) and Other (*p* = 0.001) were more likely than Democrats to be unvaccinated with a vaccinated child, while Hispanic/Latino (*p* = 0.032) respondents and those with children age 6 − 18 (*p* = 0.044) were less likely to fall in this category. In general, education and income did not predict mismatched vaccination statuses between children and adults, suggesting that this behavior is not driven by socioeconomic status.

We also asked respondents their reasons for vaccinating and not vaccinating for COVID-19 and influenza, with the option to choose more than one reason in the survey. We estimated the percentage of respondents reporting each reason for vaccinating or not vaccinating, across coarse age bands (Figure 4). Reasons for choosing not to vaccinate were varied, with the leading reasons for adults falling under “Indifference/lack of concern” (Influenza, 45% of all respondents) and “Safety concerns” (COVID-19, 45% of all respondents); no single reason was cited by the majority of respondents (*>*50%), and this heterogeneity was consistent across age (Figure 4A). The leading reason for not vaccinating a child in the home was “Safety concerns” for both COVID-19 (48%) and influenza (34%) (Figure 4A). When looking at the difference in these rates for COVID-19 versus influenza, more respondents reported “Safety concerns”, “Previous infection” or “Distrust/skepticism” for COVID-19 than influenza, while “Indifference/lack of concern” was more often cited for influenza compared to COVID-19 (Figure 4B). In contrast, vaccinated respondents were more homogeneous in their responses. Among those who were vaccinated, respondents consistently cited protecting themselves, their child, or others as a motivation, for both pathogens, across age - ranging from 75% (0-18, influenza) to 90% (60+, influenza) of respondents (Figure 4C). Health authority recommendation and work/school/travel policy were also frequently cited, with the latter decreasing with age in adults and reported more frequently for COVID-19 than for influenza. This trend is likely due to broad vaccination mandates in many workplaces and educational settings earlier in the pandemic. Older adults more often cited protecting self/child/others or health authority recommendation as reasons to vaccinate for influenza than for COVID-19, while the reverse was true for younger adults. We found similar patterns when stratifying by the overall vaccination status of adults (both vaccines, COVID-19 only, influenza only, neither vaccine) (Figure S5).

**Figure 4:**
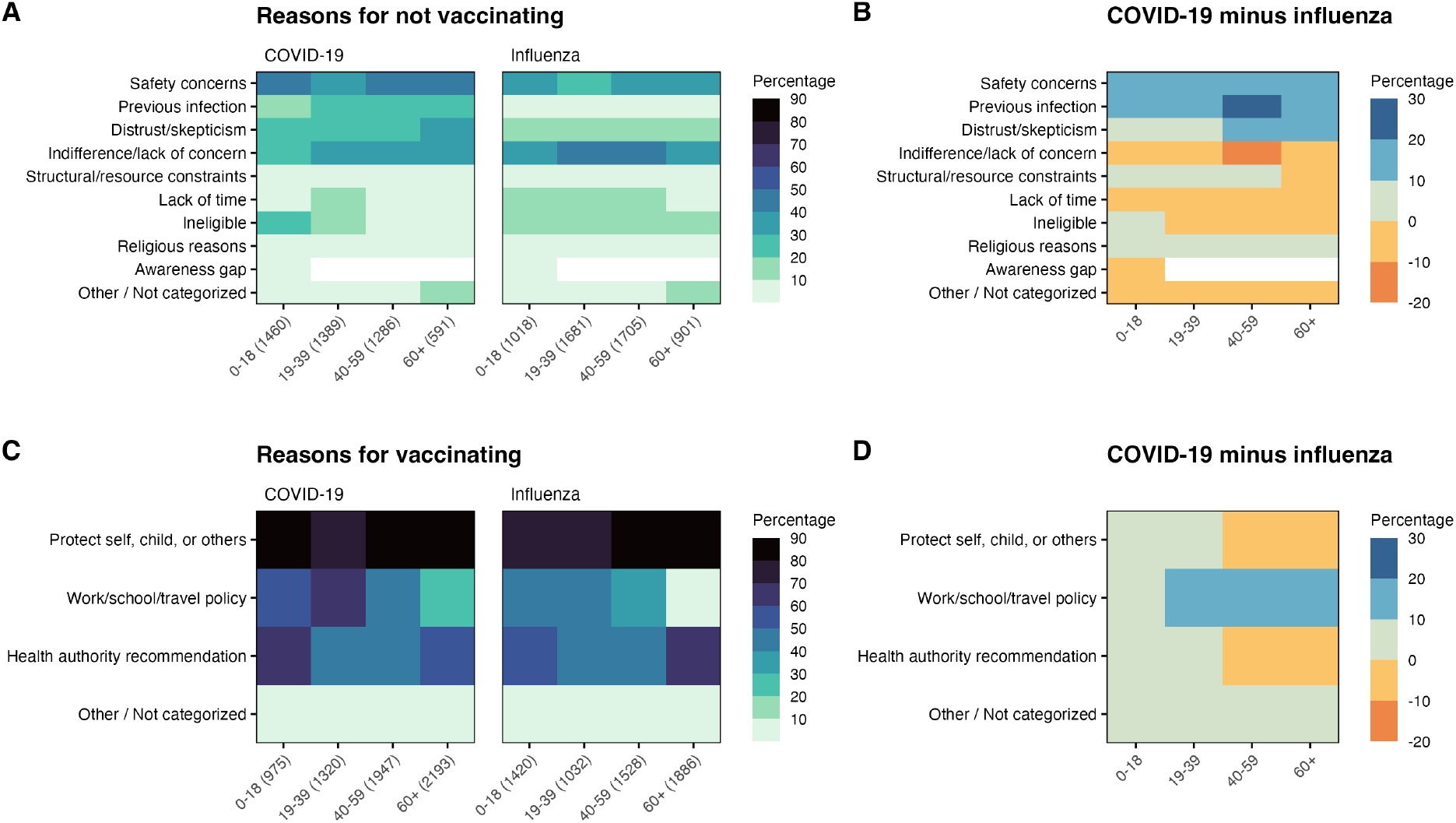
Reasons for vaccination behavior by age. Reasons reported for not being vaccinated (A) or choosing to vaccinate (C) are shown as unweighted percentages, with sample sizes in parentheses. Because participants could select more than one reason, columns will not sum to 100%. Difference in the percentage of reasons reported for COVID-19 and influenza are shown in panels B (reasons for not vaccinating) and D (reasons for vaccinating). For each age group, the total number of respondents is reported in parentheses. The grouping of specific reasons into broader categories is provided in Tables S15-S16.

Finally, having characterized the sociodemographic predictors of vaccination status (a proactive dimension of health-seeking behavior), and reasons for this decision, we sought to assess patterns in reactive behavior through analyzing the proportion seeking medical care when sick with a respiratory illness. Among 8, 860 survey respondents, 50% reported having a respiratory illness in the past 12 months, and 17% reported a respiratory illness in the past 12 months for a child in the home. We analyzed the proportion seeking medical care when sick, for the weighted samples of 4, 413 adult respondents (Table S17) and 1, 507 adults responding for children in the home (Table S18). Among adults, we observed decreasing rates of seeking care with increasing age, income, and education level, and reversed disparities by race/ethnicity and political affiliation compared to vaccination; these were all significant predictors after applying logistic regression models (Figure S6; Tables S19). We observed that adults were more likely to seek care for their children when sick compared to themselves, and this decision was predicted by political affiliation, race/ethnicity, adult age, and age of the child (Table S20). Finally, for those who chose not to seek care, we assessed the reasons, finding that the majority of respondents cited that their illness was not severe (Figure S7A). For those who did seek care, we assessed the places where care was sought (Figure S7B), finding that most respondents went to their primary care clinic or an urgent care. Taken together with Figure S6, these findings suggest that differences in care-seeking behavior reflect the distribution of illness severity, rather than access or underlying propensity to seek care.

## 4 Discussion

In this analysis of household-level survey data, we conducted a comprehensive comparison of COVID-19 versus influenza, children versus adults, and proactive versus reactive health behavior in a single population, finding substantial heterogeneity in individuals’ decisions. We find that many adults are vaccinated for one pathogen but not the other (29%), or live in households with a child whose vaccination status differs from their own (39% of adults living with children), despite CDC recommendations [27]. Against a backdrop of persistent sociodemographic disparities in vaccination across age, income, education, race/ethnicity, and political affiliation, we find consistently higher COVID-19 vaccine uptake among adults and higher influenza uptake among children. Moreover, while those choosing to vaccinate for either pathogen overwhelmingly reported wanting to protect themselves or others, reasons for not vaccinating were highly heterogeneous, ranging from distrust to structural constraints. COVID-19 vaccination remains more polarizing than influenza, with larger coverage gaps among Republicans versus Democrats and a greater share of respondents reporting safety concerns, distrust, or religious reasons for not receiving COVID-19 vaccines. Finally, we contrast proactive vaccination behavior with the reactive decision to seek medical care when sick with a respiratory illness. These results show opposite sociodemographic patterns to those observed for vaccination, which appeared primarily driven by the level of illness severity.

Lower COVID-19 than influenza vaccination rates among children are not easily attributed to differences in policy alone. During the 2024-25 season, all individuals 6 months and older were recommended by CDC to receive updated influenza and COVID-19 vaccines [27]. While other studies, including the 2025-26 National Immunization Survey [28, 29], have found lower rates of uptake for updated COVID-19 vaccines compared to influenza across all ages, we apply a different definition of COVID-19 vaccination here. We consider individuals fully vaccinated for COVID-19 if they have ever received a primary series and at least one additional dose, and the larger COVID-19 versus influenza vaccination gap among children 0-5 years may partly reflect that some children have not been alive long enough to receive both an initial COVID-19 vaccine and an updated dose. However, this cannot explain why the gap between COVID-19 and influenza coverage is larger for children of Republicans than children of Democrats, and similarly across low versus high socioeconomic status groups. Additionally, in light of our finding that work/school/travel policy and distrust/safety concerns were both cited more often for COVID-19 vaccination than for influenza, it is possible adults are more willing to take on perceived risks of COVID-19 vaccination for themselves compared to their children, particularly if externally motivated by policies in the workplace. This differential risk calculation is also supported by our descriptive finding that across demographic groups, adults sought care for their sick children more often than themselves.

Importantly, our results show that while vaccinated individuals are motivated alike, unvaccinated individuals make this decision for a range of distinct reasons, echoing the Anna Karenina Principle [30, 31] - “All happy families are alike; each unhappy family is unhappy in its own way”. This was true for both influenza and COVID-19, and persisted across age groups. Prior studies conducted before the widespread availability of COVID-19 vaccines similarly reflect a heterogeneous landscape of vaccine hesitancy towards COVID-19 or influenza vaccines [20, 32–35], with leading reasons, such as safety concerns, but limited consensus. These findings emphasize that campaigns to address low vaccine uptake cannot be one-size-fits-all, but instead must be targeted towards specific concerns and needs. For example, a recent study of influenza vaccine decision-making found that access/resource barriers were associated with higher vaccine confidence [14], indicating a need for structural intervention rather than health education in that group. Beyond quantitative assessment of vaccination motivations, qualitative work is needed in the future to better understand the distinct pathways by which vaccine hesitancy, or barriers to vaccination, develop across diverse communities (e.g. [36]).

When contextualizing vaccination behavior against seeking medical care when sick, we observed lower vaccination coverage among low socioeconomic groups, as well as Black and Hispanic respondents. In contrast, sociodemographic differences in seeking care when sick generally followed opposite trends - and our results suggest that the decision to not seek medical care depends primarily on having a mild illness. Racial/ethnic vaccination disparities observed here align with previous studies of vaccine willingness and uptake in the United States [35, 37, 38]. Higher COVID-19 vaccination rates among Asians after controlling for other demographic factors could reflect that Asians living in the United States have experienced greater coronavirus stigmatization than their white counterparts, for example being feared or treated with less respect due to coronavirus concerns [39], although our study was not designed to assess this. Importantly, wide-ranging disparities observed in this work are also likely to result from intersectional factors; after adjusting for co-variates in vaccination regression analysis, the differences seen in our descriptive results became smaller. For example, vaccination coverage gaps between political affiliation and race were less extreme, and increases in coverage across income and education levels were more gradual. This suggests that some differences were influenced by correlations in the underlying composition of demographic, socioeconomic, and political factors, rather than by a single predictor alone.

Findings from our survey should be considered in light of several limitations. First, we fielded to an online survey panel in Illinois using a quota sample rather than a gold-standard probability sample. This method was chosen because it enables collection of a larger sample than would be possible through random sampling, due to time and cost constraints. Since the time our survey was fielded, rapid developments in large language models have increased the potential for synthetic respondents in online survey panels, whose responses are consistent with assigned demographic profiles [40]. While we applied Captcha measures and took care to remove duplicate responses, LLM respondents would be very challenging to detect. Further validation of our findings can be pursued in the future through methods such as address-based sampling. Second, we fielded our survey in Illinois, and thus, further work is needed to determine if these results apply to the wider US population. Third, minority demographic groups were undersampled in some cases (Hispanic/Latino, Asian, Multiracial). To account for this issue, we weighted our sample to more closely match the population distribution of Illinois by income, age, education, and race/ethnicity. Fourth, respondents were asked to report only on the youngest child in their home; it is possible that some respondents with children 0 − 5 also have children 6 − 18, reducing our ability to observe differences between these age groups. Finally, we rely on honest and accurate reporting of behavior from respondents. However, there is a risk of recall bias - for example, respondents may remember severe illnesses more often than mild ones - or social desirability bias, for example reluctance to answer honestly about politicized behaviors like vaccination. Additionally, not all respondents answered all questions, and this missingness could be non-random, such as when reporting behavior across political affiliation. It is therefore possible that quantitative estimates of behavior could be biased. However, the qualitative consistency of our findings across observed data and regression analysis lends confidence to our results.

In summary, we found inconsistent COVID-19 and influenza vaccination behaviors across the population, with adults often making different choices for COVID-19 versus influenza, and for themselves versus their children. Sociodemographic disparities in vaccination behavior were consistently observed across age, socioeconomic status, political affiliation, and race/ethnicity, as well as across pathogens. The reasons for not vaccinating were highly heterogeneous. There were also differences in the likelihood of seeking care when sick with a respiratory illness, but these followed opposite trends to those observed for vaccination, and appeared to be driven primarily by illness severity. Our results highlight the need for strategic COVID-19 and influenza vaccination campaigns, addressing diverse barriers and hesitancies, that can reduce disease burden for all.

## 5 Methods

### Survey methods and sample selection

We conducted a statewide survey in Illinois on health and health-seeking behaviors related to respiratory pathogens in two waves: November-December 2024 (wave 1) and February-March 2025 (wave 2). We used the Qualtrics platform and respondents were recruited through Dynata, an online panel provider (Dynata, Shelton, CT, USA, https://www.dynata.com). Our survey study protocol received an IRB exemption determination from the University of Illinois Urbana-Champaign IRB (reference number IRB42-1168). All participants were provided information on the study procedure, risks, and benefits, as well as points of contact. By taking part in the study, participants confirmed that they had read the information in the consent form, had the opportunity to ask questions, and voluntarily agreed to take part in the study. We screened respondents for those over 18 and living in Illinois, and excluded those who did not meet this criteria. For both waves, we used a reCaptcha threshold of 0.5 to remove responses that were identified as likely to be bots. In Wave 2, Dynata recruited participants who had not responded in Wave 1, and we also required successful completion of a Captcha question at the beginning of the survey. Finally, we removed responses with duplicate *psid* values.

In order to conduct linear regressions controlling for key demographic factors, we selected survey respondents who provided responses to the following questions:

1. Which of the following best describes your yearly combined household income from all sources?
2. What is the highest educational level you have completed?
3. Which of the following statements best describes your current employment situation?
4. Which of the following best describes your race? (Please select all that apply.)
5. Are you of Hispanic or Latino origin (excluding origins in Spain)?
6. What is your year of birth?
7. What is your current gender identity?
8. Which political party best describes your affiliation?

We combined questions 4 and 5 into a single variable for race/ethnicity and use “Hispanic or Latino” in this work to refer to individuals of any single racial identity who report Hispanic or Latino ethnicity. Terms such as “Asian” or “White” refer to those individuals who report one racial identity and do not report Hispanic or Latino ethnicity, unless otherwise stated. Finally, we removed six respondents who reported a child under 18 in the home, but reported their child’s age as over 18 when answering questions about the child.

After these selection steps, 8, 860 respondents remained. We analyzed four distinct subsamples: adults who provided information on both their influenza and COVID-19 vaccination status (n = 8, 741), adults who provided information on their child’s influenza and COVID-19 vaccination status (n = 2, 438), adults who reported an illness in the past 12 months and provided information on care-seeking behavior for themselves (n = 4, 413), and finally adults who reported that a child in the home experienced an illness in the past 12 months, and provided information on care-seeking behavior for the child (n = 1, 507). We raked each subsample using Census data from Illinois [41] and the *weights* and *anesrake* packages in R [42, 43] with a maximum weight size of 5, to obtain representative weights by race/ethnicity, age, income, and education.

### Statistical analysis

Response variables were adult vaccination for COVID-19 or influenza, child vaccination for COVID-19 or influenza, adults seeking medical care when sick, and children seeking medical care when sick. We counted individuals as fully vaccinated for influenza if they received an immunization within the past 12 months, and fully vaccinated for COVID-19 if they had received the primary series vaccine and at least one booster (all-time). We counted individuals as seeking medical care when sick if they reported visiting a testing clinic, primary care, urgent care or emergency department, or were hospitalized. Individuals stating that they *both* did not seek care *and* engaged in one of these care seeking behaviors were excluded during sample preparation.

We used the *survey* package in R [44] and applied the *svymean()* function to estimate the weighted observed proportion engaging in vaccination or care-seeking behavior in our sample. Then, *svyciprop()* with *method = “logit”* was used to compute confidence intervals. This method estimates Wald-type 95% confidence intervals on the log-odds scale first, before transforming to the probability scale.

Next, we fit six separate quasibinomial generalized linear models (COVID-19 and influenza vaccination for adults and children; care seeking behavior for adults and children) using the *glm()* function in R, with weights identified from raking. We controlled for the following factors in all models (with reference level in parentheses): adult political affiliation (Democrat), employment (Unemployed), race/ethnicity (White), gender (Male), education (No degree/diploma), income (under $50,000 annually), and survey wave (wave 1). In adult models we controlled for respondent age, and in child models we controlled for reported age of the child.

After fitting models to data, models were used to predict vaccination and care-seeking behavior across age and key demographic features, using the *predict()* function from the *car* package in R [45]. Standard errors were first computed on the log-odds scale with the argument *se*.*fit = T*, and used to calculate Wald-type 95% confidence intervals. Then, 95% confidence intervals were transformed to the probability scale using the inverse link function.

To better understand individuals’ motivations behind each health behavior, we summarized reasons provided for behavior across age as unweighted percentages. Reasons were collapsed into broad categories which can be mapped to specific responses using Table S15. Importantly, individuals were allowed to select more than one reason for their behavior, so the percentage of respondents reporting each reason category may not sum to 100%. For individuals reporting multiple reasons within the same category, we count the category only once.

## Data Availability

Upon publication, the survey instrument as well as code and data needed to evaluate this study's conclusions and reproduce figures will be made available in a permanent repository.

## Acknowledgements

Funding for the survey, as well as support for SLL and PPM during the study, was provided by SHIELD Illinois. PPM was also supported by a grant from National Science Foundation (DMS-2436332). SLL and ASM received funding for this research from the National Institute of General Medical Sciences of the National Institutes of Health under award number R35GM156856.

## Author Contributions

Conceptualization: PPM, ASM, SLL. Funding acquisition: PPM, ASM, SLL. Project administration: SLL, PPM, ASM. Supervision: PPM, ASM, SLL. Data curation: SLL, JY. Formal analysis: SLL, JY, AA. Investigation: SLL, JY. Methodology: SLL, JY, PPM, ASM, EH, AV, LS. Software: SLL, JY. Validation: JY, SLL, AA. Visualization: PPM, SLL, JY, ASM. Writing original draft: SLL, JY. Writing - review and editing: all authors.

## Disclosure of Delegation to Generative AI

The authors declare the use of generative AI in the research and writing process. According to the GAIDeT taxonomy (2025), the following tasks were delegated to GAI tools under full human supervision: Proofread-ing and editing. The GAI tool used was: Gemini 3.5. Responsibility for the final manuscript lies entirely with the authors. GAI tools are not listed as authors and do not bear responsibility for the final outcomes.

