## Supplementary Material for "Same household, different choices: variation in health behaviors related to respiratory viruses in Illinois"

### Supplementary Materials for: Same household, different choices: variation in health behaviors related to respiratory viruses in Illinois

#### Supplementary Figures

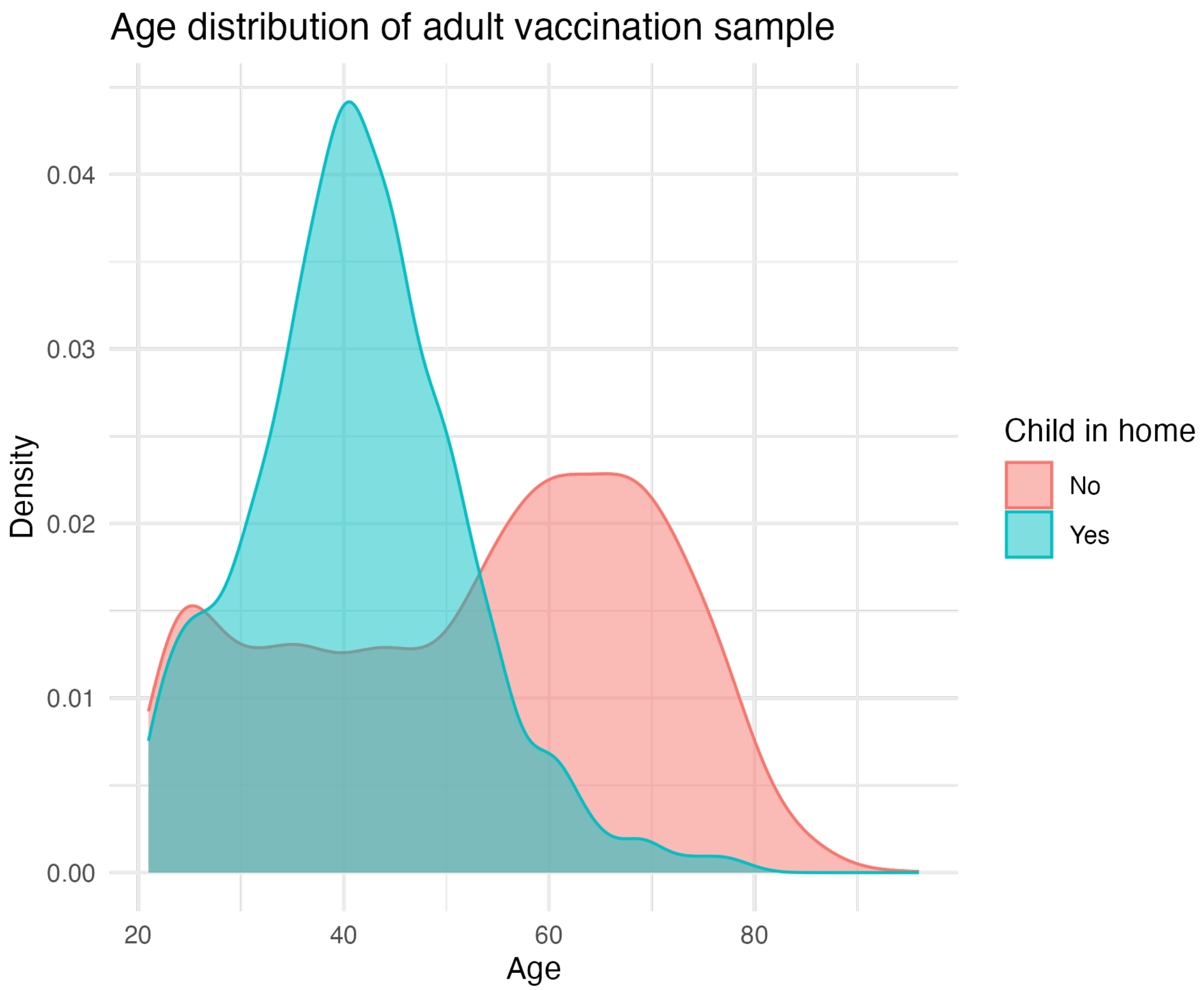


Figure S1: Density distribution of age for all adults, stratified into those who reported a child in the home (n = $2,395$) and those who did not ($n = 6,346$).

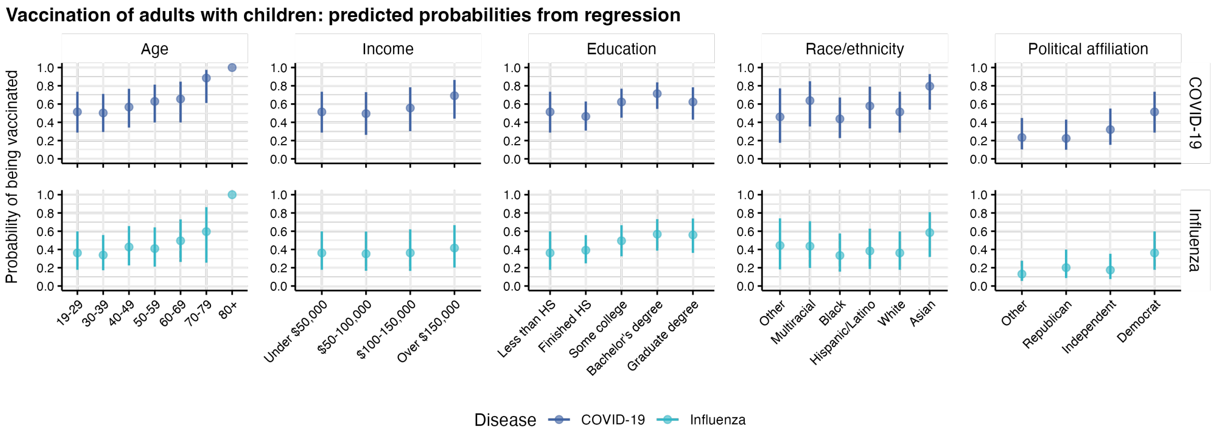


Figure S2: Predicted probability of being vaccinated as an adult reporting a child in the home, from two generalized linear models. The predicted probability is shown by age bands, income, education level, race/ethnicity, and political affiliation. 95% confidence intervals are shown for all reported values. Sample characteristics are provided in Table S8 and regression parameters in Tables S9-S10. There was only one adult observed in the age 80+ category.

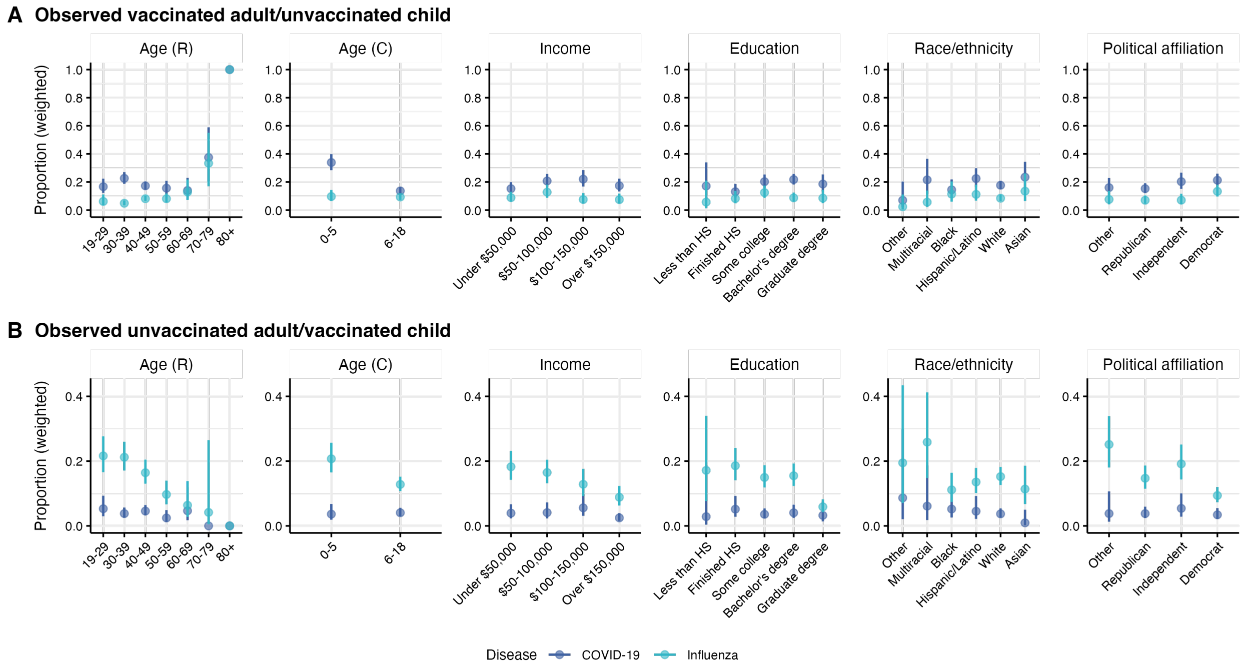


Figure S3: Observed proportion of adults who are (A) vaccinated while reporting an unvaccinated child in the home or (B) unvaccinated while reporting a vaccinated child in the home. The predicted probability is shown by respondent age bands, child age, respondent education level, income, race/ethnicity, and political affiliation. Weighted 95% confidence intervals are shown for all reported values. Sample characteristics are provided in Table S3 and regression parameters in Tables S11, S12, S13, S14. There was only one adult in the age 80+ category.

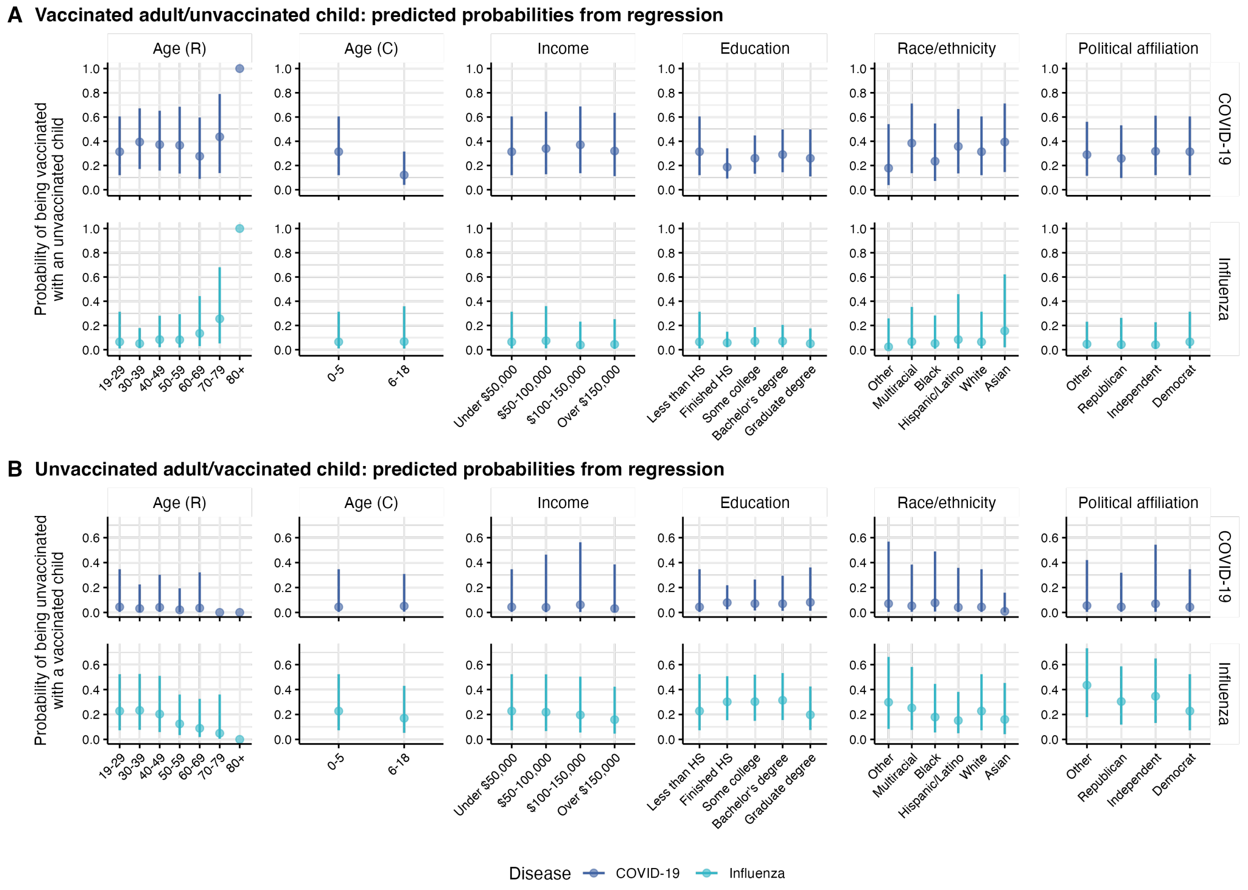


Figure S4: Predicted probability of being (A) vaccinated while reporting an unvaccinated child in the home or (B) unvaccinated while reporting a vaccinated child in the home, from four generalized linear models. The predicted probability is shown by respondent age bands, child age, respondent education level, income, race/ethnicity, and political affiliation. 95% confidence intervals are shown for all reported values. Sample characteristics are provided in Table S3 and regression parameters in Tables S11, S12, S13, S14. There was only one adult observed in the age 80+ category.

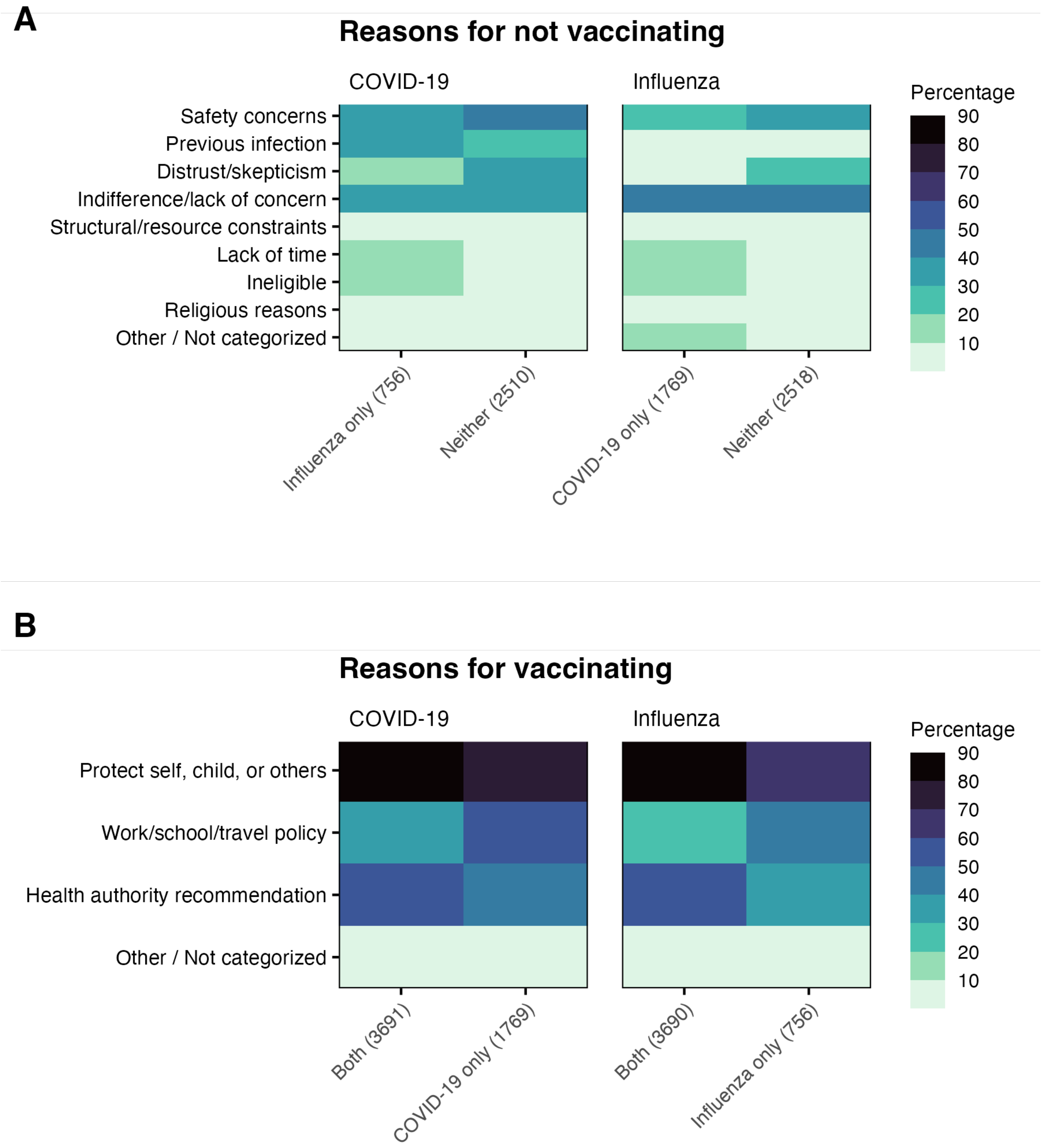


Figure S5: Reasons for adult vaccination behavior by vaccination status for both pathogens (both vaccines, COVID-19 only, influenza only, neither vaccine). Reasons reported for not being vaccinated (A) or choosing to vaccinate (B) are shown as unweighted percentages, with sample sizes in parentheses. Because participants could select more than one reason, columns will not sum to 100%. For each vaccination status, the total number of respondents is reported in parentheses.

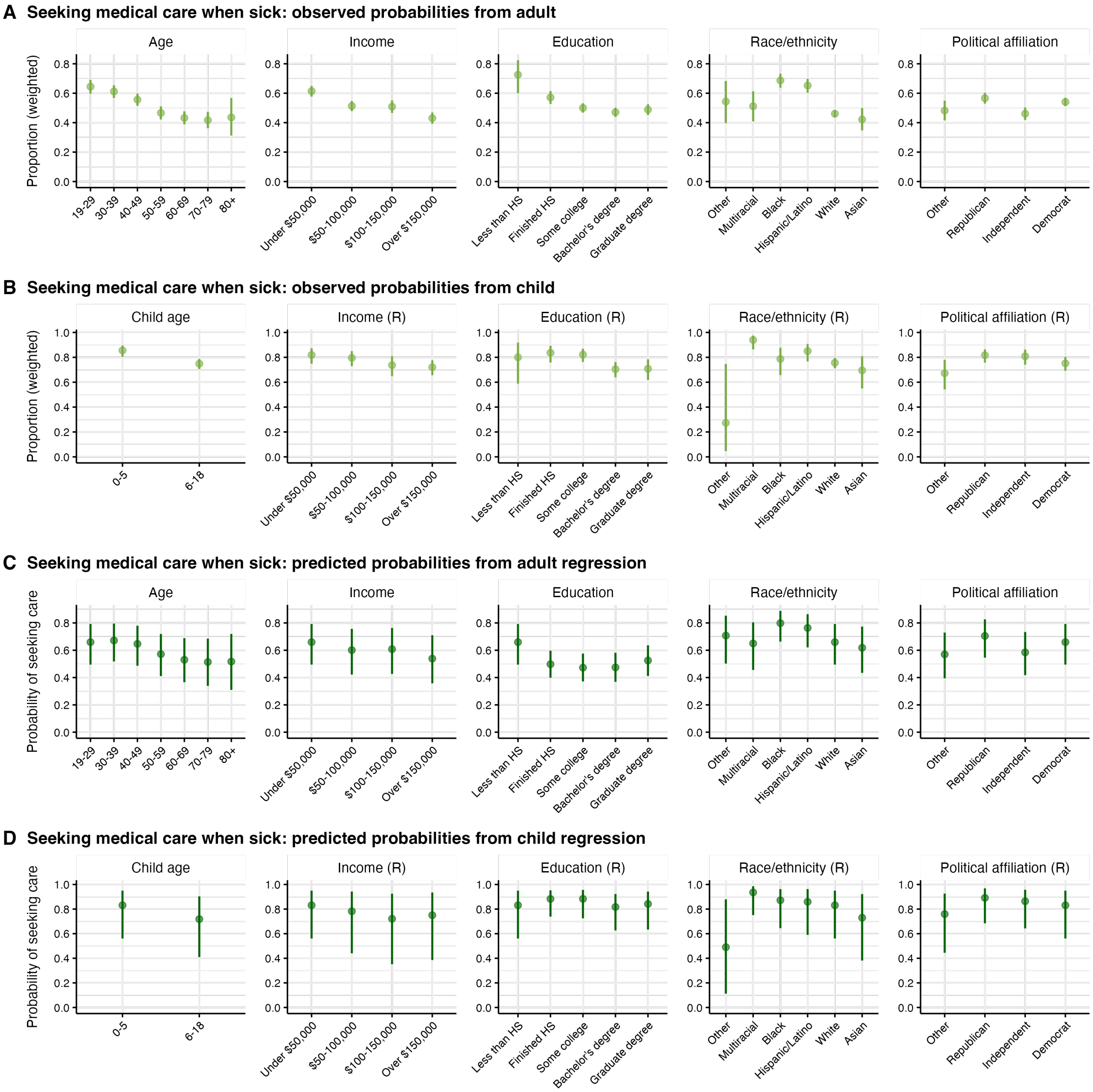


Figure S6: Proportion seeking care when sick with a respiratory illness in Illinois. (A) For adults (n = $4,418$), weighted observed proportions are shown by age, income, education level, race/ethnicity, and political affiliation. (B) For children (n = $1,513$), weighted observed proportions are shown by age band of the child,and respondent income, education, race/ethnicity, and political affiliation. (C) Predicted probability of adults seeking care when sick, from a general linear model. (D) Predicted probability of adults seeking care for a sick child in the home, from a general linear model. 95% confidence intervals are shown for all reported values. Sample characteristics are provided in Tables S17-S18.

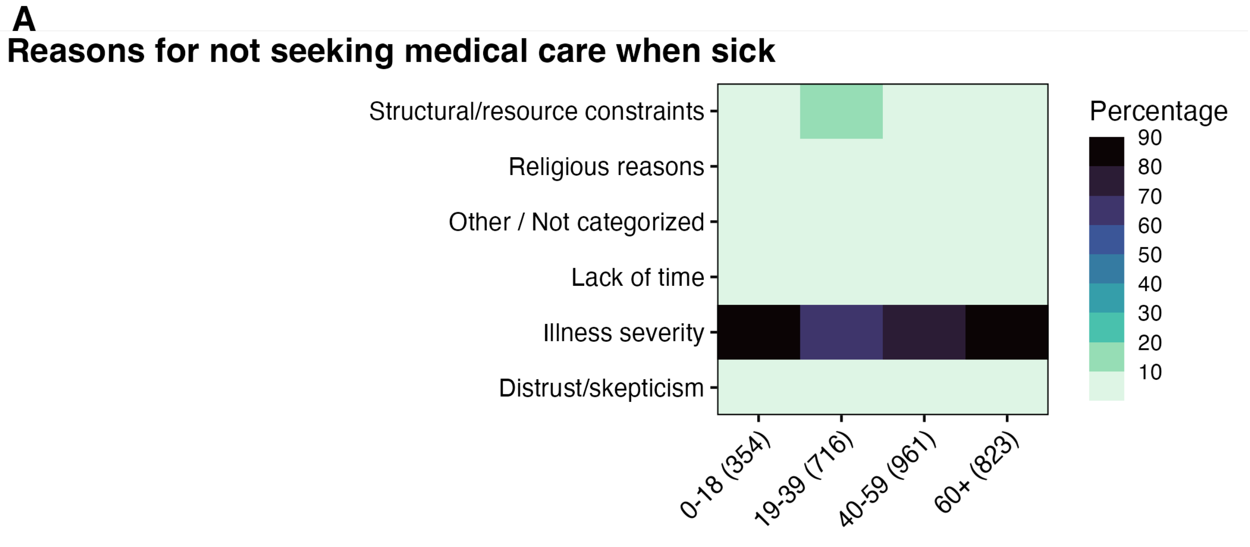


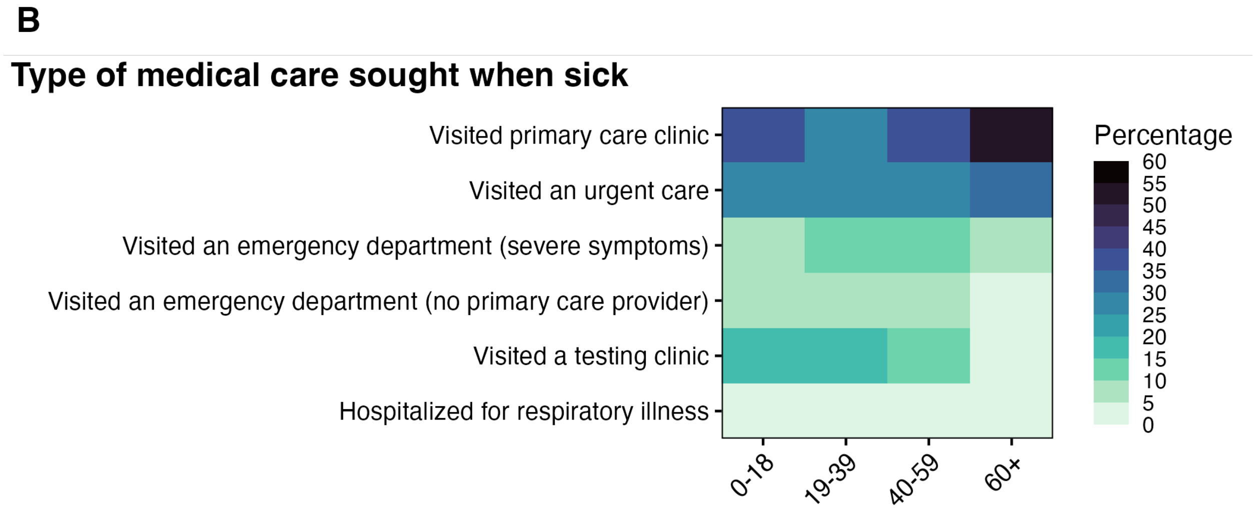


Figure S7: (A) Unweighted reasons for not seeking medical care, by age. Reasons reported are shown as unweighted percentages. Because participants could select more than one reason, columns will not sum to 100%. (B) Types of medical care sought when sick, among respondents who reported seeking medical care, as percentages. Because participants could select more than one type of care sought, columns will not sum to 100%. The number of respondents in each age group is shown in parentheses.

#### Supplementary Tables

| Sample characteristics | Weighted | Unweighted |
|---|---|---|
| Wave | | |
| 1 | 3574.94 | 3578 |
| 2 | 5166.06 | 5163 |
| Gender | | |
| Female | 5021.67 | 5143 |
| Male | 3639.07 | 3517 |
| Other | 80.26 | 81 |
| Age | | |
| 19–29 years | 1515.10 | 1188 |
| 30–39 years | 1515.19 | 1526 |
| 40–49 years | 1515.07 | 1709 |
| 50–59 years | 1398.58 | 1529 |
| 60–69 years | 1398.54 | 1548 |
| 70–79 years | 932.35 | 1068 |
| 80+ years | 466.17 | 173 |
| Income | | |
| Under $50,000 | 2756.35 | 3138 |
| $50–100,000 | 2458.64 | 2634 |
| $100–150,000 | 1583.64 | 1457 |
| Over $150,000 | 1942.37 | 1512 |
| Education | | |
| Less than a high school education | 812.77 | 164 |
| High school or equivalent | 2213.95 | 1599 |
| Some college | 2362.73 | 2699 |
| Bachelor's degree | 1995.19 | 2439 |
| Postgraduate education | 1356.37 | 1840 |
| Employment status | | |
| Permanent contract | 3285.80 | 3455 |
| Retired | 1978.28 | 1946 |
| Unemployed | 1207.92 | 1088 |
| Independent | 678.94 | 734 |
| Temporary or seasonal worker | 437.35 | 381 |
| Service provider without a specific schedule | 482.40 | 533 |
| For a fixed time or trial period | 236.94 | 235 |
| Student | 269.29 | 244 |
| I have never done paid work | 164.08 | 125 |
| Race/ethnicity: | | |
| White | 5046.86 | 5464 |
| Hispanic/Latino | 1652.23 | 1016 |
| Black or African American | 1127.77 | 1431 |
| Asian | 522.84 | 440 |
| Multiracial | 347.72 | 245 |

| Sample characteristics | Weighted | Unweighted |
|---|---|---|
| Other | 43.58 | 145 |
| Political affiliation | | |
| Republican | 2536.04 | 2417 |
| Independent | 1980.58 | 1965 |
| Democrat | 3339.46 | 3552 |
| Other | 884.91 | 807 |

Table S1: Survey sample characteristics for adult vaccination status. Weights were calculated via raking [43] with Census data [41]. Weighted sample sizes are rounded to 2 decimals.

| Sample characteristics | Weighted | Unweighted |
|---|---|---|
| Wave | | |
| 1 | 909.24 | 917 |
| 2 | 1528.76 | 1521 |
| Gender | | |
| Female | 1519.12 | 1422 |
| Male | 900.84 | 1005 |
| Other | 18.03 | 11 |
| Age | | |
| 19–29 years | 476.27 | 308 |
| 30–39 years | 476.27 | 735 |
| 40–49 years | 476.27 | 919 |
| 50–59 years | 439.63 | 358 |
| 60–69 years | 439.55 | 92 |
| 70–79 years | 125.00 | 25 |
| 80+ years | 5.00 | 1 |
| Child age | | |
| 0–5 years | 575.55 | 627 |
| 6–18 years | 1862.45 | 1811 |
| Income | | |
| Under $50,000 | 768.74 | 621 |
| $50–100,000 | 685.76 | 658 |
| $100–150,000 | 441.72 | 527 |
| Over $150,000 | 541.78 | 632 |
| Education | | |
| Less than high school | 200.00 | 40 |
| Some college | 685.27 | 633 |
| High school or equivalent | 556.44 | 354 |
| Bachelor's degree | 620.96 | 703 |
| Postgraduate education | 375.33 | 708 |
| Employment status | | |
| Permanent contract | 1089.83 | 1383 |
| Retired | 241.53 | 73 |
| Unemployed | 336.53 | 286 |
| Independent | 215.15 | 193 |
| Temporary or seasonal worker | 166.99 | 129 |
| Service provider without a specific schedule | 156.04 | 160 |
| For a fixed time or trial period | 85.99 | 107 |
| Student | 94.16 | 67 |
| I have never done paid work | 51.78 | 40 |
| Race/ethnicity | | |
| Black or African American | 300.25 | 409 |
| White | 1481.60 | 1373 |
| Hispanic/Latino | 365.42 | 416 |
| Asian | 168.05 | 133 |

| Sample characteristics | Weighted | Unweighted |
|---|---|---|
| Multiracial | 108.68 | 73 |
| Other | 14.00 | 34 |
| Political affiliation | | |
| Democrat | 886.50 | 945 |
| Independent | 452.16 | 463 |
| Republican | 828.72 | 786 |
| Other | 270.62 | 244 |

Table S2: Survey sample characteristics for child vaccination status. Weights were calculated via raking [43] with Census data [41]. Weighted sample sizes are rounded to 2 decimals.

| Sample characteristics | Weighted | Unweighted |
|---|---|---|
| Wave | | |
| 1 | 883.70 | 896 |
| 2 | 1496.30 | 1484 |
| Gender | | |
| Female | 1494.69 | 1396 |
| Male | 867.30 | 973 |
| Other | 18.01 | 11 |
| Age | | |
| 19–29 years | 465.34 | 298 |
| 30–39 years | 465.34 | 714 |
| 40–49 years | 465.34 | 903 |
| 50–59 years | 429.54 | 351 |
| 60–69 years | 429.44 | 89 |
| 70–79 years | 120.00 | 24 |
| 80+ years | 5.00 | 1 |
| Income | | |
| Under $50k | 750.45 | 596 |
| $50–100k | 669.45 | 644 |
| $100k–150k | 431.21 | 517 |
| Over $150k | 528.89 | 623 |
| Education | | |
| Less than high school | 175.00 | 35 |
| High school or equivalent | 548.75 | 344 |
| Some college | 671.63 | 624 |
| Bachelor’s degree | 617.68 | 683 |
| Postgraduate education | 366.94 | 694 |
| Employment status | | |
| Permanent contract | 1054.00 | 1351 |
| Retired | 232.93 | 71 |
| Unemployed | 331.98 | 283 |
| Independent | 215.90 | 190 |
| Temporary or seasonal worker | 165.08 | 124 |
| Service provider without a specific schedule | 150.25 | 156 |
| For a fixed time or trial period | 83.33 | 101 |
| Student | 94.12 | 65 |
| I have never done paid work | 52.42 | 39 |
| Race/ethnicity | | |
| Black or African American | 296.91 | 390 |
| Asian | 165.54 | 129 |
| White | 1444.53 | 1349 |
| Multiracial | 105.32 | 73 |
| Hispanic/Latino | 353.90 | 406 |
| Other | 13.79 | 33 |

| Sample characteristics | Weighted | Unweighted |
|---|---|---|
| Political affiliation | | |
| Democrat | 877.39 | 926 |
| Republican | 805.57 | 763 |
| Independent | 440.97 | 453 |
| Other | 256.08 | 238 |

Table S3: Survey sample characteristics for adult versus child vaccination status. Weights were calculated via raking [43] with Census data [41]. Weighted sample sizes are rounded to 2 decimals.

| Variable | Estimate | p-value |
|---|---|---|
| **Political affiliation** | | |
| Democrat | – | – |
| Independent | -0.75 [-0.92, -0.58] | $< 0.001$ |
| Republican | -1.38 [-1.55, -1.22] | $< 0.001$ |
| Other | -0.95 [-1.17, -0.72] | $< 0.001$ |
| **Employment** | | |
| Unemployed | – | – |
| For a fixed time or trial period | 0.33 [-0.04, 0.71] | 0.084 |
| I have never done paid work | -0.07 [-0.75, 0.61] | 0.838 |
| Independent | -0.12 [-0.4, 0.16] | 0.397 |
| Permanent contract | 0.53 [0.32, 0.75] | $< 0.001$ |
| Retired | 0.6 [0.29, 0.91] | $< 0.001$ |
| Service provider without a specific schedule | -0.06 [-0.33, 0.22] | 0.696 |
| Student | 0.52 [0.11, 0.92] | 0.014 |
| Temporary or seasonal worker | 0.23 [-0.13, 0.59] | 0.204 |
| **Race/ethnicity** | | |
| White | – | – |
| Asian | 0.97 [0.65, 1.29] | $< 0.001$ |
| Black or African American | -0.2 [-0.41, 0.02] | 0.069 |
| Hispanic/Latino | 0.12 [-0.07, 0.31] | 0.211 |
| Multiracial | 0.18 [-0.22, 0.58] | 0.375 |
| Other | -0.55 [-1.02, -0.07] | 0.023 |
| **Gender** | | |
| Male | – | – |
| Female | -0.03 [-0.16, 0.10] | 0.686 |
| Other | 0.32 [-0.50, 1.13] | 0.450 |
| **Education** | | |
| No degree/diploma | – | – |
| High school | 0.14 [-0.22, 0.51] | 0.444 |
| Some college | 0.43 [0.06, 0.80] | 0.021 |
| Bachelors | 1.04 [0.661, 1.416] | $< 0.001$ |
| Postgrad | 1.11 [0.72, 1.5] | $< 0.001$ |
| **Age** | | |
| 19–29 years | – | – |
| 30–39 years | -0.05 [-0.27, 0.17] | 0.659 |
| 40–49 years | 0.33 [0.11, 0.56] | 0.004 |
| 50–59 years | 0.67 [0.43, 0.91] | $< 0.001$ |
| 60–69 years | 1.12 [0.84, 1.4] | $< 0.001$ |
| 70–79 years | 1.73 [1.37, 2.09] | $< 0.001$ |
| 80+ years | 2.16 [1.51, 2.81] | $< 0.001$ |
| **Income** | | |
| Less than $50,000 | – | – |
| $50–100,000 | -0.02 [-0.17, 0.14] | 0.848 |

| Variable | Estimate | p-value |
|---|---|---|
| $100–150,000 | 0.15 [-0.05, 0.35] | 0.130 |
| Over $150,000 | 0.42 [0.20, 0.64] | $< 0.001$ |
| **Wave** | | |
| 1 | – | – |
| 2 | 0.00 [-0.12, 0.13] | 0.94 |

Table S4: Adjusted logistic regression estimates for adult COVID-19 vaccination. Table reports coefficient estimates and 95% confidence intervals for demographic, socioeconomic, and political predictors, along with p-values.

| Variable | Estimate | p-value |
|---|---|---|
| **Political affiliation** | | |
| Democrat | – | – |
| Independent | -0.77 [-0.92, -0.62] | $< 0.001$ |
| Other | -1.1 [-1.32, -0.89] | $< 0.001$ |
| Republican | -0.82 [-0.97, -0.67] | $< 0.001$ |
| **Employment** | | |
| Unemployed | – | – |
| For a fixed time or trial period | 0.25 [-0.12, 0.62] | 0.186 |
| I have never done paid work | 0.21 [-0.40, 0.81] | 0.505 |
| Independent | -0.19 [-0.46, 0.08] | 0.169 |
| Permanent contract | 0.31 [0.09, 0.52] | 0.005 |
| Retired | 0.57 [0.30, 0.84] | $< 0.001$ |
| Service provider without a specific schedule | 0.10 [-0.18, 0.38] | 0.469 |
| Student | 0.34 [-0.08, 0.76] | 0.114 |
| Temporary or seasonal worker | 0.24 [-0.10, 0.59] | 0.164 |
| **Race/ethnicity** | | |
| White | – | – |
| Asian | 0.48 [0.24, 0.73] | $< 0.001$ |
| Black or African American | -0.05 [-0.24, 0.14] | 0.616 |
| Hispanic/Latino | 0.13 [-0.05, 0.31] | 0.147 |
| Multiracial | 0.20 [-0.12, 0.52] | 0.227 |
| Other | -0.17 [-0.77, 0.44] | 0.583 |
| **Gender** | | |
| Male | – | – |
| Female | 0.06 [-0.06, 0.18] | 0.342 |
| Other | 0.13 [-0.51, 0.76] | 0.691 |
| **Education** | | |
| No degree/diploma | – | – |
| High school | -0.05 [-0.40, 0.31] | 0.799 |
| Some college | 0.24 [-0.11, 0.60] | 0.173 |
| Bachelors | 0.62 [0.25, 0.98] | 0.001 |
| Postgrad | 0.96 [0.59, 1.34] | 0 |
| **Age** | | |
| 19-29 years | – | – |
| 30–39 years | -0.02 [-0.24, 0.20] | 0.879 |
| 40–49 years | 0.29 [0.07, 0.51] | 0.009 |
| 50–59 years | 0.33 [0.10, 0.55] | 0.004 |
| 60–69 years | 0.78 [0.54, 1.03] | $< 0.001$ |
| 70–79 years | 1.35 [1.044, 1.648] | $< 0.001$ |
| 80+ years | 1.69 [1.20, 2.19] | $< 0.001$ |
| **Income** | | |
| Less than $50,000 | – | – |
| $50–100,000 | 0.05 [-0.10, 0.20] | 0.513 |

| Variable | Estimate | p-value |
|---|---|---|
| $100–150,000 | 0.23 [0.04, 0.43] | 0.017 |
| Over $150,000 | 0.27 [0.06, 0.47] | 0.011 |
| **Wave** | | |
| 1 | – | – |
| 2 | 0.08 [-0.03, 0.20] | 0.159 |

Table S5: Adjusted logistic regression estimates for adult influenza vaccination. Table reports coefficient estimates and 95% confidence intervals for demographic, socioeconomic, and political predictors, along with p-values.

| Variable | Estimate | p-value |
|---|---|---|
| **Political affiliation** | | |
| Democrat | – | – |
| Independent | -0.64 [-1.02, -0.26] | 0.001 |
| Other | -1.06 [-1.62, -0.50] | $< 0.001$ |
| Republican | -1.09 [-1.42, -0.77] | $< 0.001$ |
| **Employment** | | |
| Unemployed | – | – |
| For a fixed time or trial period | -0.18 [-1.17, 0.82] | 0.729 |
| I have never done paid work | -0.52 [-1.53, 0.49] | 0.311 |
| Independent | 0.4 [-0.21, 1.01] | 0.195 |
| Permanent contract | -0.04 [-0.54, 0.45] | 0.869 |
| Retired | -0.61 [-1.39, 0.17] | 0.126 |
| Service provider without a specific schedule | -0.44 [-1.12, 0.25] | 0.209 |
| Student | 0.22 [-0.62, 1.07] | 0.603 |
| Temporary or seasonal worker | 0.09 [-0.60, 0.78] | 0.797 |
| **Race/ethnicity** | | |
| White | – | – |
| Asian | 0.68 [0.18, 1.18] | 0.008 |
| Black or African American | 0.12 [-0.39, 0.62] | 0.653 |
| Hispanic/Latino | 0.16 [-0.23, 0.55] | 0.425 |
| Multiracial | 0.35 [-0.39, 1.09] | 0.357 |
| Other | 0.65 [-0.48, 1.79] | 0.26 |
| **Gender** | | |
| Male | – | – |
| Female | -0.45 [-0.74, -0.16] | 0.002 |
| Other | 0.80 [-0.48, 2.08] | 0.221 |
| **Education** | | |
| No degree/diploma | – | – |
| High school | 0.37 [-0.56, 1.3] | 0.433 |
| Some college | 0.8 [-0.10, 1.71] | 0.082 |
| Bachelors | 1.05 [0.10, 2.00] | 0.031 |
| Postgrad | 0.91 [-0.06, 1.88] | 0.066 |
| **Child age** | | |
| 0–5 years | – | – |
| 6–18 years | 1.34 [0.97, 1.71] | $< 0.001$ |
| **Income** | | |
| $50,000 | – | – |
| $50–100,000 | -0.16 [-0.58, 0.26] | 0.451 |
| $100–150,000 | 0.11 [-0.38, 0.59] | 0.666 |
| Over $150,000 | 0.57 [0.07, 1.08] | 0.027 |
| **Parent Age (continuous)** | | |
| Age | 0.01 [0.00, 0.03] | 0.033 |
| **Wave** | | |

| Variable | Estimate | p-value |
|---|---|---|
| 1 | – | – |
| 2 | 0.13 [-0.15, 0.41] | 0.372 |

Table S6: Adjusted logistic regression estimates for child COVID-19 vaccination. Table reports coefficient estimates and 95% confidence intervals for demographic, socioeconomic, and political predictors, along with p-values.

| Variable | Estimate | p-value |
|---|---|---|
| **Political affiliation** | | |
| Democrat | – | – |
| Independent | -0.49 [-0.87, -0.11] | 0.011 |
| Other | -0.51 [-0.98, -0.05] | 0.031 |
| Republican | -0.48 [-0.80, -0.15] | 0.004 |
| **Employment** | | |
| Unemployed | – | – |
| For a fixed time or trial period | -0.58 [-1.28, 0.11] | 0.102 |
| I have never done paid work | -0.27 [-1.34, 0.80] | 0.618 |
| Independent | -0.16 [-0.72, 0.41] | 0.588 |
| Permanent contract | -0.17 [-0.60, 0.26] | 0.434 |
| Retired | -0.27 [-1.01, 0.46] | 0.463 |
| Service provider without a specific schedule | -0.15 [-0.75, 0.44] | 0.61 |
| Student | 0.28 [-0.50, 1.06] | 0.478 |
| Temporary or seasonal worker | 0.27 [-0.37, 0.90] | 0.409 |
| **Race/ethnicity** | | |
| White | – | – |
| Asian | 0.25 [-0.27, 0.77] | 0.354 |
| Black or African American | -0.15 [-0.62, 0.31] | 0.515 |
| Hispanic/Latino | -0.28 [-0.64, 0.08] | 0.131 |
| Multiracial | 0.37 [-0.30, 1.04] | 0.28 |
| Other | 0.68 [-0.18, 1.55] | 0.12 |
| **Gender** | | |
| Male | – | – |
| Female | -0.48 [-0.76, -0.20] | 0.001 |
| Other | -0.84 [-2.67, 0.99] | 0.368 |
| **Education** | | |
| No degree/diploma | – | – |
| High school | 0.04 [-0.66, 0.75] | 0.906 |
| Some college | 0.40 [-0.31, 1.1] | 0.269 |
| Bachelors | 0.74 [0.00, 1.47] | 0.049 |
| Postgrad | 0.62 [-0.17, 1.40] | 0.123 |
| **Child age** | | |
| 0–5 years | – | – |
| 6–18 years | -0.18 [-0.47, 0.11] | 0.23 |
| **Income** | | |
| Less than $50,000 | – | – |
| $50–100,000 | -0.11 [-0.48, 0.25] | 0.548 |
| $100–150,000 | 0.05 [-0.40, 0.5] | 0.822 |
| Over $150,000 | 0.20 [-0.27, 0.66] | 0.408 |
| **Parent age (continuous)** | | |
| Age | -0.01 [-0.02, 0.00] | 0.150 |
| **Wave** | | |

| Variable | Estimate | p-value |
|---|---|---|
| 1 | – | – |
| 2 | 0.39 [0.12, 0.66] | 0.004 |

Table S7: Adjusted logistic regression estimates for child influenza vaccination. Table reports coefficient estimates and 95% confidence intervals for demographic, socioeconomic, and political predictors, along with p-values.

| Sample characteristics | Weighted | Unweighted |
|---|---|---|
| Wave | | |
| 1 | 885.59 | 899 |
| 2 | 1509.41 | 1496 |
| Gender | | |
| Female | 1500.96 | 1405 |
| Male | 876.06 | 979 |
| Other | 17.98 | 11 |
| Age | | |
| 19–29 years | 467.40 | 300 |
| 30–39 years | 467.40 | 719 |
| 40–49 years | 467.40 | 905 |
| 50–59 years | 431.45 | 355 |
| 60–69 years | 431.35 | 90 |
| 70–79 years | 125.00 | 25 |
| 80+ years | 5.00 | 1 |
| Income | | |
| Under $50,000 | 755.18 | 602 |
| $50–100,000 | 673.67 | 647 |
| $100–150,000 | 433.93 | 519 |
| Over $150,000 | 532.22 | 627 |
| Education | | |
| No degree | 175.00 | 35 |
| High school | 552.29 | 347 |
| Some college | 673.36 | 629 |
| Bachelor's degree | 621.53 | 686 |
| Postgraduate education | 372.82 | 698 |
| Employment status | | |
| Permanent contract | 1058.61 | 1356 |
| Retired | 237.65 | 72 |
| Unemployed | 333.37 | 287 |
| Independent | 218.23 | 191 |
| Temporary or seasonal worker | 165.90 | 125 |
| Service provider without a specific schedule | 151.15 | 158 |
| For a fixed time or trial period | 83.98 | 102 |
| Student | 93.77 | 65 |
| I have never done paid work | 52.35 | 39 |
| Race/ethnicity | | |
| Black or African American | 300.89 | 393 |
| Asian | 166.32 | 129 |
| White | 1452.33 | 1357 |
| Multiracial | 105.77 | 73 |
| Hispanic/Latino | 355.83 | 409 |
| Other | 13.86 | 34 |

| Sample characteristics | Weighted | Unweighted |
|---|---|---|
| Political affiliation | | |
| Democrat | 885.69 | 931 |
| Republican | 806.65 | 767 |
| Independent | 446.59 | 458 |
| Other | 256.06 | 239 |

Table S8: Survey sample characteristics for adults with a child in the household, reporting their own vaccination status. Weights were calculated via raking [43] with Census data [41]. Weighted sample sizes are rounded to 2 decimals.

| Variable | Estimate | p-value |
|---|---|---|
| **Political affiliation** | | |
| Democrat | – | – |
| Independent | -0.81 [-1.17, -0.44] | $< 0.001$ |
| Other | -1.24 [-1.71, -0.78] | $< 0.001$ |
| Republican | -1.30 [-1.64, -0.96] | $< 0.001$ |
| **Employment** | | |
| Unemployed | – | – |
| For a fixed time or trial period | -0.07 [-0.83, 0.68] | 0.850 |
| I have never done paid work | -0.68 [-1.71, 0.34] | 0.192 |
| Independent | 0.26 [-0.31, 0.83] | 0.375 |
| Permanent contract | 0.19 [-0.27, 0.64] | 0.416 |
| Retired | -0.12 [-1.02, 0.78] | 0.794 |
| Service provider without a specific schedule | -0.81 [-1.46, -0.17] | 0.013 |
| Student | 0.22 [-0.59, 1.03] | 0.590 |
| Temporary or seasonal worker | 0.28 [-0.41, 0.97] | 0.430 |
| **Race/ethnicity** | | |
| White | – | – |
| Asian | 1.3 [0.633, 1.97] | $< 0.001$ |
| Black or African American | -0.31 [-0.75, 0.13] | 0.170 |
| Hispanic/Latino | 0.26 [-0.13, 0.65] | 0.192 |
| Multiracial | 0.51 [-0.21, 1.23] | 0.164 |
| Other | -0.22 [-1.23, 0.80] | 0.673 |
| **Gender** | | |
| Male | – | – |
| Female | -0.35 [-0.63, -0.07] | 0.015 |
| Other | 0.8 [-1.09, 2.70] | 0.407 |
| **Education** | | |
| No degree/diploma | – | – |
| High school | -0.20 [-0.97, 0.58] | 0.618 |
| Some college | 0.44 [-0.31, 1.20] | 0.251 |
| Bachelors | 0.86 [0.06, 1.65] | 0.035 |
| Postgrad | 0.45 [-0.40, 1.29] | 0.302 |
| **Age** | | |
| 19–29 years | – | – |
| 30–39 years | -0.04 [-0.43, 0.34] | 0.821 |
| 40–49 years | 0.21 [-0.18, 0.61] | 0.289 |
| 50–59 years | 0.47 [0.03, 0.92] | 0.039 |
| 60–69 years | 0.59 [-0.05, 1.23] | 0.072 |
| 70–79 years | 1.98 [0.66, 3.31] | 0.003 |
| 80+ years | 13.96 [11.75, 16.17] | $< 0.001$ |
| **Income** | | |
| Less than $50,000 | – | – |
| $50–100,000 | -0.07 [-0.46, 0.31] | 0.703 |

| Variable | Estimate | p-value |
|---|---|---|
| $100–150,000 | 0.17 [-0.29, 0.64] | 0.472 |
| Over $150,000 | 0.75 [0.28, 1.22] | 0.002 |
| **Wave** | | |
| 1 | – | – |
| 2 | 0.07 [-0.21, 0.35] | 0.617 |

Table S9: Adjusted logistic regression estimates for adult COVID-19 vaccination status, conditional on having a child in the household. Table reports coefficient estimates and 95% confidence intervals for demographic, socioeconomic, and political predictors, along with p-values.

| Variable | Estimate | p-value |
|---|---|---|
| **Political affiliation** | | |
| Democrat | – | – |
| Independent | -0.99 [-1.35, -0.62] | $< 0.001$ |
| Other | -1.33 [-1.81, -0.85] | $< 0.001$ |
| Republican | -0.81 [-1.14, -0.49] | $< 0.001$ |
| **Employment** | | |
| Unemployed | – | – |
| For a fixed time or trial period | -0.12 [-0.87, 0.63] | 0.753 |
| I have never done paid work | -0.13 [-1.46, 1.20] | 0.848 |
| Independent | -0.06 [-0.61, 0.50] | 0.841 |
| Permanent contract | 0.11 [-0.33, 0.55] | 0.630 |
| Retired | 0.24 [-0.61, 1.10] | 0.576 |
| Service provider without a specific schedule | -0.2 [-0.77, 0.37] | 0.496 |
| Student | 0.60 [-0.22, 1.41] | 0.153 |
| Temporary or seasonal worker | 0.23 [-0.41, 0.87] | 0.478 |
| **Race/ethnicity** | | |
| White | – | – |
| Asian | 0.91 [0.403, 1.414] | $< 0.001$ |
| Black or African American | -0.12 [-0.58, 0.34] | 0.605 |
| Hispanic/Latino | 0.09 [-0.30, 0.49] | 0.642 |
| Multiracial | 0.31 [-0.36, 0.98] | 0.366 |
| Other | 0.34 [-0.52, 1.20] | 0.438 |
| **Gender** | | |
| Male | – | – |
| Female | -0.12 [-0.40, 0.16] | 0.400 |
| Other | -0.93 [-2.76, 0.89] | 0.317 |
| **Education** | | |
| No degree/diploma | – | – |
| High school | 0.13 [-0.68, 0.94] | 0.758 |
| Some college | 0.55 [-0.25, 1.35] | 0.181 |
| Bachelors | 0.84 [0.02, 1.66] | 0.046 |
| Postgrad | 0.81 [-0.07, 1.68] | 0.071 |
| **Age** | | |
| 19–29 years | – | – |
| 30–39 years | -0.10 [-0.49, 0.29] | 0.615 |
| 40–49 years | 0.27 [-0.12, 0.67] | 0.178 |
| 50–59 years | 0.20 [-0.23, 0.63] | 0.360 |
| 60–69 years | 0.55 [-0.05, 1.14] | 0.072 |
| 70–79 years | 0.95 [-0.24, 2.15] | 0.118 |
| 80+ years | 13.80 [11.62, 15.98] | $< 0.001$ |
| **Income** | | |
| $50,000 | – | – |
| $50–100,000 | -0.04 [-0.41, 0.33] | 0.826 |

| Variable | Estimate | p-value |
|---|---|---|
| $100–150,000 | 0.00 [-0.46, 0.47] | 0.989 |
| Over $150,000 | 0.23 [-0.25, 0.71] | 0.356 |
| **Wave** | | |
| 1 | – | – |
| 2 | 0.34 [0.07, 0.61] | 0.014 |

Table S10: Adjusted logistic regression estimates for adult influenza vaccination status, given that they have a child in the home. Table reports coefficient estimates and 95% confidence intervals for demographic, socioeconomic, and political predictors, along with p-values.

| Variable | Estimate | p-value |
|---|---|---|
| **Political affiliation** | | |
| Democrat | – | – |
| Independent | 0.02 [-0.43, 0.46] | 0.946 |
| Other | -0.12 [-0.68, 0.45] | 0.687 |
| Republican | -0.27 [-0.66, 0.12] | 0.168 |
| **Employment** | | |
| Unemployed | – | – |
| For a fixed time or trial period | 0.71 [-0.10, 1.52] | 0.086 |
| I have never done paid work | -0.94 [-2.19, 0.30] | 0.138 |
| Independent | -0.2 [-0.91, 0.52] | 0.591 |
| Permanent contract | 0.44 [-0.12, 1.01] | 0.125 |
| Retired | 1.27 [0.29, 2.26] | 0.012 |
| Service provider without a specific schedule | -0.84 [-1.65, -0.03] | 0.041 |
| Student | 0.13 [-0.74, 1.01] | 0.764 |
| Temporary or seasonal worker | 0.63 [-0.11, 1.37] | 0.095 |
| **Race/ethnicity** | | |
| White | – | – |
| Asian | 0.35 [-0.20, 0.90] | 0.208 |
| Black or African American | -0.40 [-1.08, 0.29] | 0.256 |
| Hispanic/Latino | 0.20 [-0.25, 0.65] | 0.387 |
| Multiracial | 0.31 [-0.43, 1.06] | 0.411 |
| Other | -0.74 [-1.94, 0.45] | 0.223 |
| **Gender** | | |
| Male | – | – |
| Female | -0.12 [-0.44, 0.20] | 0.463 |
| Other | 0.54 [-1.54, 2.63] | 0.610 |
| **Education** | | |
| No degree/diploma | – | – |
| High school | -0.69 [-1.69, 0.31] | 0.179 |
| Some college | -0.26 [-1.27, 0.74] | 0.606 |
| Bachelors | -0.11 [-1.14, 0.92] | 0.832 |
| Postgrad | -0.27 [-1.39, 0.86] | 0.644 |
| **Age** | | |
| 19–29 years | – | – |
| 30–39 years | 0.35 [-0.10, 0.81] | 0.130 |
| 40–49 years | 0.26 [-0.22, 0.74] | 0.286 |
| 50–59 years | 0.24 [-0.35, 0.82] | 0.430 |
| 60–69 years | -0.18 [-0.96, 0.61] | 0.655 |
| 70–79 years | 0.53 [-0.63, 1.68] | 0.371 |
| 80+ years | 14.54 [12.26, 16.81] | $< 0.001$ |
| **Child age** | | |
| 0–5 years | – | – |
| 6–18 years | -1.20 [-1.56, -0.83] | $< 0.001$ |

| Variable | Estimate | p-value |
|---|---|---|
| **Income** | | |
| Less than $50,000 | – | – |
| $50–100,000 | 0.12 [-0.37, 0.61] | 0.635 |
| $100–150,000 | 0.25 [-0.30, 0.81] | 0.369 |
| Over $150,000 | 0.03 [-0.62, 0.67] | 0.936 |
| **Wave** | | |
| 1 | – | – |
| 2 | -0.14 [-0.48, 0.19] | 0.394 |

Table S11: Adjusted logistic regression estimates for adults vaccinated for COVID-19, with unvaccinated children. Table reports coefficient estimates and 95% confidence intervals for demographic, socioeconomic, and political predictors, along with p-values.

| Variable | Estimate | p-value |
|---|---|---|
| **Political affiliation** | | |
| Democrat | – | – |
| Independent | -0.48 [-1.17, 0.20] | 0.167 |
| Other | -0.35 [-1.06, 0.36] | 0.339 |
| Republican | -0.43 [-0.98, 0.13] | 0.133 |
| **Employment** | | |
| Unemployed | – | – |
| For a fixed time or trial period | 0.73 [-0.80, 2.26] | 0.351 |
| I have never done paid work | -1.04 [-3.25, 1.17] | 0.357 |
| Independent | -0.95 [-2.20, 0.29] | 0.134 |
| Permanent contract | 0.43 [-0.34, 1.20] | 0.273 |
| Retired | 0.81 [-0.25, 1.88] | 0.135 |
| Service provider without a specific schedule | 0.49 [-0.68, 1.66] | 0.410 |
| Student | -0.25 [-1.68, 1.19] | 0.737 |
| Temporary or seasonal worker | 0.05 [-1.28, 1.38] | 0.942 |
| **Race/ethnicity** | | |
| White | – | – |
| Asian | 0.98 [0.10, 1.86] | 0.030 |
| Black or African American | -0.24 [-1.03, 0.55] | 0.552 |
| Hispanic/Latino | 0.28 [-0.42, 0.98] | 0.434 |
| Multiracial | 0.04 [-1.01, 1.09] | 0.947 |
| Other | -1.06 [-2.84, 0.73] | 0.247 |
| **Gender** | | |
| Male | – | – |
| Female | 0.13 [-0.36, 0.61] | 0.607 |
| Other | -0.25 [-2.34, 1.84] | 0.813 |
| **Education** | | |
| No degree/diploma | – | – |
| High school | -0.12 [-1.62, 1.37] | 0.872 |
| Some college | 0.09 [-1.30, 1.49] | 0.895 |
| Bachelors | 0.09 [-1.32, 1.50] | 0.903 |
| Postgrad | -0.26 [-1.76, 1.24] | 0.734 |
| **Age** | | |
| 19–29 years | – | – |
| 30–39 years | -0.26 [-1.08, 0.56] | 0.538 |
| 40–49 years | 0.28 [-0.53, 1.09] | 0.502 |
| 50–59 years | 0.26 [-0.60, 1.11] | 0.557 |
| 60–69 years | 0.81 [-0.25, 1.88] | 0.135 |
| 70–79 years | 1.60 [0.30, 2.90] | 0.016 |
| 80+ years | 15.97 [13.483, 18.455] | $< 0.001$ |
| **Child age** | | |
| 0–5 years | – | – |
| 6–18 years | 0.04 [-0.52, 0.60] | 0.886 |

| Variable | Estimate | p-value |
|---|---|---|
| **Income** | | |
| Less than $50,000 | – | – |
| $50–100,000 | 0.15 [-0.49, 0.78] | 0.648 |
| $100–150,000 | -0.52 [-1.31, 0.27] | 0.198 |
| Over $150,000 | -0.4 [-1.17, 0.38] | 0.316 |
| **Wave** | | |
| 1 | – | – |
| 2 | -0.22 [-0.69, 0.25] | 0.353 |

Table S12: Adjusted logistic regression estimates for adults vaccinated for influenza, with unvaccinated children. Table reports coefficient estimates and 95% confidence intervals for demographic, socioeconomic, and political predictors, along with p-values

| Variable | Estimate | p-value |
|---|---|---|
| **Political affiliation** | | |
| Democrat | – | – |
| Independent | 0.48 [-0.26, 1.23] | 0.203 |
| Other | 0.23 [-1.01, 1.47] | 0.716 |
| Republican | 0.03 [-0.61, 0.67] | 0.924 |
| **Employment** | | |
| Unemployed | – | – |
| For a fixed time or trial period | 1.05 [-0.37, 2.48] | 0.148 |
| I have never done paid work | -2.00 [-4.30, 0.30] | 0.088 |
| Independent | -0.15 [-1.34, 1.05] | 0.809 |
| Permanent contract | 0.03 [-0.92, 0.98] | 0.958 |
| Retired | 0.42 [-1.66, 2.50] | 0.694 |
| Service provider without a specific schedule | 0.26 [-0.98, 1.49] | 0.685 |
| Student | -0.96 [-2.74, 0.81] | 0.288 |
| Temporary or seasonal worker | 0.56 [-0.78, 1.89] | 0.413 |
| **Race/ethnicity** | | |
| White | – | – |
| Asian | -1.59 [-3.43, 0.26] | 0.092 |
| Black or African American | 0.59 [-0.28, 1.47] | 0.182 |
| Hispanic/Latino | -0.06 [-0.82, 0.69] | 0.87 |
| Multiracial | 0.19 [-1.14, 1.52] | 0.78 |
| Other | 0.50 [-1.19, 2.19] | 0.562 |
| **Gender** | | |
| Male | – | – |
| Female | -1.00 [-1.73, -0.27] | 0.008 |
| Other | 0.54 [-1.50, 2.59] | 0.603 |
| **Education** | | |
| No degree/diploma | – | – |
| High school | 0.62 [-1.56, 2.80] | 0.578 |
| Some college | 0.50 [-1.63, 2.64] | 0.644 |
| Bachelors | 0.49 [-1.96, 2.94] | 0.697 |
| Postgrad | 0.65 [-1.76, 3.07] | 0.595 |
| **Age** | | |
| 19–29 years | – | – |
| 30–39 years | -0.35 [-1.11, 0.41] | 0.366 |
| 40–49 years | -0.07 [-0.80, 0.66] | 0.850 |
| 50–59 years | -0.74 [-1.78, 0.29] | 0.158 |
| 60–69 years | -0.23 [-1.45, 1.00] | 0.719 |
| 70–79 years | -15.79 [-17.49, -14.09] | $< 0.001$ |
| 80+ years | -15.36 [-18.241, -12.478] | $< 0.001$ |
| **Child age** | | |
| 0–5 years | – | – |
| 6–18 years | 0.15 [-0.53, 0.84] | 0.658 |

| Variable | Estimate | p-value |
|---|---|---|
| **Income** | | |
| Less than \$50,000 | – | – |
| \$50–100,000 | -0.07 [-1.13, 0.98] | 0.892 |
| \$100–150,000 | 0.35 [-0.80, 1.50] | 0.550 |
| Over \$150,000 | -0.36 [-1.52, 0.81] | 0.550 |
| **Wave** | | |
| 1 | – | – |
| 2 | -0.36 [-1.00, 0.29] | 0.278 |

Table S13: Adjusted logistic regression estimates for adults unvaccinated for COVID-19, with vaccinated children. Table reports coefficient estimates and 95% confidence intervals for demographic, socioeconomic, and political predictors, along with p-values.

| Variable | Estimate | p-value |
|---|---|---|
| **Political affiliation** | | |
| Democrat | – | – |
| Independent | 0.59 [0.12, 1.06] | 0.013 |
| Other | 0.96 [0.40, 1.52] | 0.001 |
| Republican | 0.39 [-0.06, 0.85] | 0.090 |
| **Employment** | | |
| Unemployed | – | – |
| For a fixed time or trial period | -0.26 [-1.22, 0.70] | 0.592 |
| I have never done paid work | -0.48 [-1.72, 0.76] | 0.451 |
| Independent | -0.50 [-1.21, 0.20] | 0.161 |
| Permanent contract | -0.14 [-0.67, 0.39] | 0.603 |
| Retired | 0.33 [-0.82, 1.48] | 0.570 |
| Service provider without a specific schedule | 0.44 [-0.26, 1.13] | 0.217 |
| Student | -0.37 [-1.17, 0.44] | 0.370 |
| Temporary or seasonal worker | 0.25 [-0.55, 1.05] | 0.541 |
| **Race/ethnicity** | | |
| White | – | – |
| Asian | -0.44 [-1.10, 0.23] | 0.198 |
| Black or African American | -0.30 [-0.83, 0.23] | 0.269 |
| Hispanic/Latino | -0.50 [-0.96, -0.04] | 0.032 |
| Multiracial | 0.14 [-0.62, 0.90] | 0.725 |
| Other | 0.37 [-0.53, 1.27] | 0.426 |
| **Gender** | | |
| Male | – | – |
| Female | -0.50 [-0.86, -0.15] | 0.005 |
| Other | -0.14 [-1.82, 1.55] | 0.874 |
| **Education** | | |
| No degree/diploma | – | – |
| High school | 0.38 [-0.56, 1.33] | 0.428 |
| Some college | 0.39 [-0.56, 1.34] | 0.419 |
| Bachelors | 0.44 [-0.57, 1.45] | 0.393 |
| Postgrad | -0.18 [-1.27, 0.91] | 0.744 |
| **Age** | | |
| 19–29 years | – | – |
| 30–39 years | 0.03 [-0.42, 0.48] | 0.899 |
| 40–49 years | -0.14 [-0.62, 0.33] | 0.556 |
| 50–59 years | -0.73 [-1.31, -0.14] | 0.015 |
| 60–69 years | -1.10 [-2.09, -0.11] | 0.029 |
| 70–79 years | -1.74 [-4.00, 0.53] | 0.133 |
| 80+ years | -12.97 [-15.36, -10.59] | $< 0.001$ |
| **Child age** | | |
| 0–5 years | – | – |
| 6–18 years | -0.36 [-0.72, -0.01] | 0.044 |

| Variable | Estimate | p-value |
|---|---|---|
| **Income** | | |
| Less than $50,000 | – | – |
| $50–100,000 | -0.05 [-0.50, 0.40] | 0.823 |
| $100–150,000 | -0.19 [-0.79, 0.41] | 0.541 |
| Over $150,000 | -0.45 [-1.11, 0.21] | 0.185 |
| **Wave** | | |
| 1 | – | – |
| 2 | -0.05 [-0.40, 0.30] | 0.780 |

Table S14: Adjusted logistic regression estimates for adults unvaccinated for influenza, with vaccinated children. Table reports coefficient estimates and 95% confidence intervals for demographic, socioeconomic, and political predictors, along with p-values.

| Reason category | Specific reason |
|---|---|
| Health authority recommendation | Because it was recommended by my healthcare provider<br>Because it was recommended by my child's healthcare provider<br>Because it was recommended by public officials |
| Protect self, child, or others | To protect myself and others<br>To protect my child and others<br>To protect my unborn child<br>I am concerned about infecting others<br>To protect the child and others |
| Work/school/travel policy | For job related reasons<br>For school related reasons<br>For travel related reasons<br>Policy at my school<br>Policy at my workplace<br>Policy at my child's school |
| Distrust/skepticism | Distrust of pharmaceutical companies<br>Distrust of the government<br>Distrust of medical providers<br>I don't believe in vaccines<br>I don't believe in mainstream medicine |
| Previous infection | I have already been infected<br>My child has already been infected<br>Your child has already been infected<br>They have already been infected |
| Safety concerns | Belief that the vaccine is riskier than infection<br>Concerns about getting it through the vaccine<br>Concerns about getting ill through the vaccine<br>I am worried about adverse effects<br>Belief that the vaccine is riskier than an infection |
| Religious reasons | Religious reasons |
| Lack of time | Lack of time<br>Unable to take time off of work<br>I did not have time<br>Unable to take time off of school<br>I was unable to take time off of school to keep them home<br>I was unable to take time off of work to keep them home |
| Structural/resource constraints | Lack of money<br>Lack of transportation<br>I did not have transportation to the testing site<br>I live in crowded housing conditions<br>I could not afford it<br>I am afraid of losing my job |
| Awareness gap | I was not aware of the vaccine<br>I was not aware of the vaccine and/or booster |

| | |
|---|---|
| | I was not aware of RSV preventive antibody |
| Ineligible | My child is too young or not eligible<br>I have experienced adverse effects<br>My child has experienced adverse effects |
| Hasn't been sick<br>Illness severity<br><br><br><br><br><br>Indifference/lack of concern | I have not been sick<br>I was severely ill<br>The illness was not severe<br>My child was only mildly ill<br>I was not severely ill<br>My child's illness was severe<br>My illness was not severe<br>I prefer not to know what illness I have |
| | I did not think it was necessary<br>I am not concerned about infection<br>Respiratory illnesses are not serious illnesses<br>Belief that COVID-19 is not a serious illness<br>Belief that influenza is not a serious illness |
| Other / Not categorized | – |

Table S15: Classification of responses to reasons for health-seeking behavior into broad categories.

| Question / Response | Count |
|---|---|
| Adult sought medical care for child's respiratory illness | |
| Hospitalized for respiratory illness | 58 |
| Visited a testing clinic | 310 |
| Visited an urgent care | 452 |
| Visited their primary care clinic | 708 |
| Visited an emergency department due to severe symptoms | 167 |
| Visited an emergency department due to lack of a primary care provider | 116 |
| Did not seek medical care | 356 |
| Child vaccinated against COVID-19 | |
| Yes | 1524 |
| No | 973 |
| Child vaccinated against influenza | |
| Yes | 1459 |
| No | 1044 |
| Type of COVID-19 vaccine received by child | |
| Primary series only | 528 |
| Primary series with one booster | 631 |
| Primary series with multiple boosters | 361 |
| Reasons child was not vaccinated against COVID-19 | |
| Concern about adverse effects | 539 |
| Concern about getting infection through the vaccine | 153 |
| Not concerned about infection | 377 |
| Belief vaccine is riskier than infection | 294 |
| Distrust of pharmaceutical companies | 197 |
| Distrust of the government | 185 |
| Do not believe in vaccines | 78 |
| Not aware of vaccine and/or booster | 106 |
| Child already infected | 285 |
| Child experienced adverse effects | 101 |
| Child too young or not eligible | 217 |
| Lack of time | 105 |
| Lack of money | 61 |
| Lack of transportation | 39 |
| Religious reasons | 69 |
| Other | 97 |
| Reasons child was not vaccinated against influenza | |
| Not concerned about infection | 325 |
| Concern about adverse effects | 217 |
| Concern about getting infection through the vaccine | 91 |
| Belief vaccine is riskier than infection | 181 |
| Distrust of pharmaceutical companies | 100 |
| Distrust of the government | 89 |
| Do not believe in vaccines | 58 |

| Question / Response | Count |
|---|---|
| Not aware of the vaccine | 75 |
| Child too young or not eligible | 100 |
| Child already infected | 68 |
| Child experienced adverse effects | 75 |
| Lack of time | 118 |
| Lack of money | 32 |
| Lack of transportation | 19 |
| Religious reasons | 37 |
| Other | 86 |
| Reasons for not seeking medical care for child | |
| Illness was not severe | 331 |
| Lack of money | 14 |
| Lack of time | 13 |
| Lack of transportation | 8 |
| Distrust of medical providers | 4 |
| Do not believe in mainstream medicine | 2 |
| Religious reasons | 5 |
| Other | 19 |
| Reasons child was vaccinated against COVID-19 | |
| To protect the child and others | 832 |
| Recommended by healthcare provider | 567 |
| Recommended by public officials | 352 |
| School-related requirements | 518 |
| Other | 21 |
| Reasons child was vaccinated against influenza | |
| To protect the child and others | 1101 |
| Recommended by healthcare provider | 801 |
| Recommended by public officials | 312 |
| School-related requirements | 706 |
| Other | 21 |
| Adult sought medical care for respiratory illness | |
| Did not seek medical care | 1906 |
| Visited a testing clinic | 352 |
| Visited primary care clinic | 973 |
| Visited an urgent care | 705 |
| Visited emergency department due to severe symptoms | 236 |
| Visited emergency department due to lack of a primary care provider | 135 |
| Hospitalized for respiratory illness | 73 |
| Adult vaccinated against COVID-19 | |
| Yes | 7237 |
| No | 1771 |
| Adult vaccinated against influenza | |
| Yes | 4543 |

| Question / Response | Count |
|---|---|
| No | 4382 |
| Type of COVID-19 vaccine received by adult | |
| Primary series only | 1627 |
| Primary series with one booster | 2263 |
| Primary series with multiple boosters | 3338 |
| Reasons adult did not seek medical care | |
| Illness was not severe | 1741 |
| Lack of money | 160 |
| Lack of time | 134 |
| Lack of transportation | 49 |
| Distrust of medical providers | 33 |
| Do not believe in mainstream medicine | 20 |
| Religious reasons | 5 |
| Other | 153 |
| Reasons adult was vaccinated against COVID-19 | |
| To protect myself and others | 4642 |
| Recommended by healthcare provider | 2296 |
| Recommended by public officials | 1819 |
| Job-related requirements | 1802 |
| Travel-related requirements | 875 |
| School-related requirements | 479 |
| To protect an unborn child | 195 |
| Other | 172 |
| Reasons adult was vaccinated against influenza | |
| To protect myself and others | 3764 |
| Recommended by healthcare provider | 2173 |
| Recommended by public officials | 1007 |
| Job-related requirements | 1059 |
| School-related requirements | 376 |
| To protect an unborn child | 223 |
| Other | 112 |

Table S16: Raw response counts for survey questions on vaccination status and medical care–seeking behavior among adults and children. Follow-up questions on reasons were asked conditionally based on respondents' prior responses.

| Sample characteristics | Weighted | Unweighted |
|---|---|---|
| Wave | | |
| 1 | 1699.83 | 1741 |
| 2 | 2713.17 | 2672 |
| Gender | | |
| Female | 2657.08 | 2715 |
| Male | 1703.81 | 1641 |
| Other | 52.11 | 57 |
| Age | | |
| 19–29 years | 764.92 | 658 |
| 30–39 years | 764.92 | 797 |
| 40–49 years | 764.92 | 953 |
| 50–59 years | 706.08 | 754 |
| 60–69 years | 706.08 | 743 |
| 70–79 years | 470.72 | 446 |
| 80+ years | 235.36 | 62 |
| Income | | |
| Under $50,000 | 1391.49 | 1353 |
| $50–100,000 | 1241.29 | 1352 |
| $100–150,000 | 799.55 | 817 |
| Over $150,000 | 980.67 | 891 |
| Education | | |
| less than high school | 310.00 | 62 |
| High school or equivalent | 1145.76 | 613 |
| Some college | 1222.75 | 1318 |
| Bachelor's degree | 1032.54 | 1337 |
| Postgraduate education | 701.95 | 1083 |
| Employment status | | |
| Permanent contract | 1686.26 | 1920 |
| Retired | 1024.83 | 872 |
| Unemployed | 529.80 | 466 |
| Independent | 323.23 | 353 |
| Temporary or seasonal worker | 223.33 | 192 |
| Service provider without a specific schedule | 270.64 | 307 |
| For a fixed time or trial period | 140.80 | 135 |
| Student | 151.73 | 127 |
| I have never done paid work | 62.38 | 41 |
| Race/ethnicity: | | |
| White | 2554.14 | 2869 |
| Hispanic/Latino | 835.70 | 538 |
| Asian | 268.70 | 214 |
| Multiracial | 172.67 | 146 |
| Black or African American | 559.24 | 584 |
| Other | 22.55 | 62 |

| Sample characteristics | Weighted | Unweighted |
|---|---|---|
| Political affiliation | | |
| Independent | 930.14 | 932 |
| Democrat | 1872.60 | 1943 |
| Republican | 1217.44 | 1171 |
| Other | 392.82 | 367 |

Table S17: Survey sample characteristics for adults seeking medical care when sick with a respiratory illness. Weights were calculated via raking [43] with Census data [41]. Weighted sample sizes are rounded to 2 decimals.

| Sample characteristics | Weighted | Unweighted |
|---|---|---|
| Wave | | |
| 1 | 563.46 | 555 |
| 2 | 943.54 | 952 |
| Gender | | |
| Female | 954.08 | 904 |
| Male | 545.06 | 599 |
| Other | 7.86 | 4 |
| Age | | |
| 19–29 years | 310.21 | 183 |
| 30–39 years | 310.21 | 456 |
| 40–49 years | 310.21 | 611 |
| 50–59 years | 286.35 | 199 |
| 60–69 years | 215.00 | 43 |
| 70–79 years | 70.00 | 14 |
| 80+ years | 5.00 | 1 |
| Child age | | |
| 0–5 years | 382.37 | 423 |
| 6–18 years | 1124.63 | 1084 |
| Income | | |
| Under $50,000 | 475.18 | 338 |
| $50–100,000 | 423.89 | 389 |
| $100–150,000 | 273.04 | 348 |
| Over $150,000 | 334.89 | 432 |
| Education | | |
| Less than high school | 125.00 | 25 |
| High school or equivalent | 340.55 | 185 |
| Some college | 422.72 | 362 |
| Bachelor’s degree | 385.07 | 456 |
| Postgraduate education | 233.66 | 479 |
| Employment status | | |
| Permanent contract | 685.49 | 880 |
| Retired | 147.11 | 41 |
| Unemployed | 205.11 | 165 |
| Independent | 125.07 | 110 |
| Temporary or seasonal worker | 108.63 | 81 |
| Service provider without a specific schedule | 105.93 | 105 |
| For a fixed time or trial period | 59.12 | 70 |
| Student | 48.80 | 36 |
| I have never done paid work | 21.73 | 19 |
| Race/ethnicity | | |
| Asian | 106.49 | 85 |
| White | 918.79 | 903 |
| Multiracial | 69.24 | 48 |
| Black or African American | 178.85 | 196 |

| Sample characteristics | Weighted | Unweighted |
|---|---|---|
| Hispanic/Latino | 226.14 | 257 |
| Other | 7.49 | 18 |
| Political affiliation | | |
| Republican | 488.05 | 486 |
| Democrat | 562.59 | 592 |
| Independent | 297.77 | 289 |
| Other | 158.59 | 140 |

Table S18: Survey sample characteristics for adults seeking medical care for their child when sick with a respiratory illness. Weights were calculated via raking [43] with Census data [41]. Weighted sample sizes are rounded to 2 decimals.

| Variable | Estimate | p-value |
|---|---|---|
| **Political affiliation** | | |
| Democrat | – | – |
| Independent | -0.32 [-0.53, -0.11] | 0.003 |
| Other | -0.38 [-0.68, -0.08] | 0.014 |
| Republican | 0.21 [0.01, 0.41] | 0.040 |
| **Employment** | | |
| Unemployed | – | – |
| For a fixed time or trial period | 0.98 [0.41, 1.54] | 0.001 |
| I have never done paid work | 0.71 [-0.12, 1.54] | 0.093 |
| Independent | 0.21 [-0.17, 0.59] | 0.275 |
| Permanent contract | 0.21 [-0.08, 0.51] | 0.157 |
| Retired | 0.26 [-0.12, 0.64] | 0.184 |
| Service provider without a specific schedule | 0.26 [-0.18, 0.70] | 0.247 |
| Student | 0.17 [-0.40, 0.74] | 0.561 |
| Temporary or seasonal worker | 0.86 [0.40, 1.33] | $< 0.001$ |
| **Race/ethnicity** | | |
| White | – | – |
| Asian | -0.18 [-0.53, 0.17] | 0.312 |
| Black or African American | 0.72 [0.45, 0.98] | $< 0.001$ |
| Hispanic/Latino | 0.51 [0.27, 0.75] | $< 0.001$ |
| Multiracial | -0.04 [-0.47, 0.39] | 0.847 |
| Other | 0.22 [-0.35, 0.80] | 0.448 |
| **Gender** | | |
| Male | – | – |
| Female | -0.01 [-0.18, 0.17] | 0.948 |
| Other | 0.08 [-0.58, 0.74] | 0.816 |
| **Education** | | |
| No degree/diploma | – | – |
| High school | -0.67 [-1.27, -0.07] | 0.029 |
| Some college | -0.77 [-1.37, -0.17] | 0.011 |
| Bachelors | -0.76 [-1.38, -0.15] | 0.015 |
| Postgrad | -0.56 [-1.18, 0.07] | 0.079 |
| **Age** | | |
| 19–29 years | – | – |
| 30–39 years | 0.06 [-0.24, 0.35] | 0.717 |
| 40–49 years | -0.06 [-0.36, 0.249] | 0.721 |
| 50–59 years | -0.37 [-0.67, -0.07] | 0.016 |
| 60–69 years | -0.54 [-0.87, -0.21] | 0.002 |
| 70–79 years | -0.61 [-1.02, -0.19] | 0.004 |
| 80+ years | -0.59 [-1.23, 0.05] | 0.069 |
| **Income** | | |
| Less than $50,000 annually | – | – |
| $50–100,000 | -0.25 [-0.48, -0.02] | 0.032 |

| Variable | Estimate | p-value |
|---|---|---|
| $100–150,000 | -0.22 [-0.48, 0.04] | 0.096 |
| Over $150,000 | -0.51 [-0.79, -0.22] | $<$ 0.001 |
| **Wave** | | |
| 1 | – | – |
| 2 | 0.31 [0.147, 0.475] | 0 |

Table S19: Adjusted logistic regression estimates for whether adults seek medical care during respiratory illnesses. Table reports coefficient estimates and 95% confidence intervals for demographic, socioeconomic, and political predictors, along with p-values.

| Variable | Estimate | p-value |
|---|---|---|
| **Political affiliation** | | |
| Democrat | – | – |
| Independent | 0.25 [-0.27, 0.78] | 0.343 |
| Other | -0.44 [-1.08, 0.19] | 0.169 |
| Republican | 0.51 [0.01, 1.00] | 0.045 |
| **Employment** | | |
| Unemployed | – | – |
| For a fixed time or trial period | -0.11 [-1.28, 1.07] | 0.859 |
| I have never done paid work | 0.19 [-1.29, 1.67] | 0.800 |
| Independent | 1.05 [0.18, 1.91] | 0.018 |
| Permanent contract | 0.85 [0.22, 1.47] | 0.008 |
| Retired | 1.21 [0.19, 2.23] | 0.020 |
| Service provider without a specific schedule | 0.19 [-0.75, 1.14] | 0.687 |
| Student | 0.55 [-0.76, 1.86] | 0.411 |
| Temporary or seasonal worker | 2.37 [1.13, 3.61] | $< 0.001$ |
| **Race/ethnicity** | | |
| White | – | – |
| Asian | -0.6 [-1.30, 0.10] | 0.092 |
| Black or African American | 0.32 [-0.37, 1.02] | 0.365 |
| Hispanic/Latino | 0.22 [-0.40, 0.83] | 0.488 |
| Multiracial | 1.08 [0.07, 2.09] | 0.037 |
| Other | -1.63 [-3.18, -0.09] | 0.038 |
| **Gender** | | |
| Male | – | – |
| Female | -0.47 [-0.94, 0.00] | 0.050 |
| Other | 11.91 [10.607, 13.214] | $< 0.001$ |
| **Education** | | |
| No degree/diploma | – | – |
| High school | 0.43 [-0.65, 1.50] | 0.437 |
| Some college | 0.43 [-0.64, 1.50] | 0.431 |
| Bachelors | -0.10 [-1.19, 1.00] | 0.860 |
| Postgrad | 0.08 [-1.08, 1.24] | 0.894 |
| **Child age** | | |
| 0–5 years | – | – |
| 6–18 years | -0.66 [-1.13, -0.19] | 0.006 |
| **Income** | | |
| Less than $50,000 | – | – |
| $50–100,000 | -0.31 [-0.96, 0.33] | 0.337 |
| $100–150,000 | -0.64 [-1.33, 0.04] | 0.065 |
| Over $150,000 | -0.49 [-1.18, 0.19] | 0.159 |
| **Parent age (continuous)** | | |
| Age | -0.02 [-0.04, 0.00] | 0.029 |
| **Wave** | | |

| Variable | Estimate | p-value |
|---|---|---|
| 1 | – | – |
| 2 | 0.48 [0.10, 0.86] | 0.014 |

Table S20: Adjusted logistic regression estimates for whether adults seek medical care for children in the household during respiratory illnesses. Table reports coefficient estimates and 95% confidence intervals for demographic, socioeconomic, and political predictors, along with p-values.
